# Multi-strain modeling of influenza vaccine effectiveness in older adults and its dependence on antigenic distance

**DOI:** 10.1101/2024.05.03.24306809

**Authors:** Séverine Urdy, Matthias Hanke, Ana I. Toledo, Nicolas Ratto, Evgueni Jacob, Emmanuel Peyronnet, Jean-Baptiste Gourlet, Sandra S. Chaves, Edward Thommes, Laurent Coudeville, Jean-Pierre Boissel, Eulalie Courcelles, Lara Bruezière

## Abstract

Influenza vaccine effectiveness (VE) varies seasonally due to host, virus and vaccine characteristics. To investigate how antigenic matching and dosage impact VE, we developed a mechanistic knowledge-based mathematical model. Immunization with a split vaccine is modeled for exposure to A/H1N1 or A/H3N2 virus strains. The model accounts for cross-reactivity of immune cells elicited during previous immunizations with new antigens. We simulated vaccine effectiveness (sVE) of high dose (HD) versus standard dose (SD) vaccines in the older population, from 2011 to 2022. We find that sVE is highly dependent on antigenic matching and that higher dosage improves immunogenicity, activation and memory formation of immune cells. Across all simulations, the HD vaccine performs better than the SD vaccine, supporting the use of the HD vaccine in the older population. This model could be adapted to predict the impact of alternative virus strain selection on clinical outcomes in future influenza seasons.

## Introduction

Measuring the full impact of influenza is difficult due to its varied clinical manifestations [1]. In the U.S., influenza is estimated to have caused 9 - 41 million illnesses, 140,000 - 710,000 hospitalizations, and 12,000 - 52,000 deaths annually from 2010 to 2020 [2]. Seasonal disease burden and severity vary, influenced by population immunity and how well vaccine strains match circulating viruses [3–4].

The immune system is split into an innate component, responding quickly but non-specifically to pathogens, and an adaptive component, responding slowly but specifically [5]. The adaptive immune system includes B and T cells (mediating respectively humoral and cellular responses), targeting short peptide fragments (epitopes) on infected cells and antigen-presenting cells (APCs), neutralizing viruses and destroying infected cells via cytolysis. Only specific B and T cells are stimulated, clonally expanded and maintained long-term.

Influenza vaccines aim to elicit antibodies, mainly against hemagglutinin (HA), but also neuraminidase (NA) [5]. Antigenic drift arises from mutations in these immunodominant epitopes creating new strains which can evade previously established immunity [3,6]. Seasonal vaccine effectiveness (VE) can vary widely due to antigenic mismatches between vaccine and circulating strains [3,5,7–8]. Predicting which strains will dominate in the next season remains a challenge, complicating decisions on vaccine strain selection [4]. Additionally, interpreting serological data is difficult due to unknown patient exposure histories to antigenically related cross-reactive and strains [4].

We focus on the widely-used inactivated split vaccines. These are made from egg-grown viruses, which can lead to issues with egg adaptation where the vaccine strain acquires key mutations that improve proliferation in eggs, but can decrease the match to the circulating viral strain. [9–11].

Quantitative modeling is essential to understand the fluctuating VE due to the complex interaction between antigenic drift and patient immunity. Within-host models, especially for influenza A, have been crucial for simulating immune responses to viral infections [12–19]. Few models address vaccine immunogenicity [19–22], particularly for influenza [23–24]. To our knowledge, this multi-strain model is unique in predicting population-level VE from within-host models for virtual patients with varied immune backgrounds. It compares the efficacy of different split vaccine doses in older patients, incorporating antigenic distances (AgD) between historical, vaccine, and circulating strains for deeper insights.

## Results

### Our model is built from literature-derived knowledge

The main model assumptions about viral dynamics, vaccine pharmacokinetics and interactions between antigens and cellular behaviors are illustrated in Figure 1. One key assumption is that specific memory cells and antibodies elicited against one viral strain during previous infection or vaccination cross-react with a newly encountered strain as long as the two strains are antigenically similar [7,25–27]. Immunization leads to the formation, maturation and maintenance of strain-specific antibodies and immune cells in response to antigen exposure. The immunization process resulting from infection or vaccination starts when APCs are exposed to antigen in tissues (muscle, lung or lymph nodes) and present phagocytosed antigens to other cells (Fig. 1A, D). APCs then migrate to secondary lymphoid tissues, where they prime and activate naïve B and T cells [28]. CD4+ T cells further enhance the activation of primed B cells, which differentiate into B cells producing non-specific antibodies, and later differentiate into memory B cells secreting strain-specific antibodies (Fig. 1C, E). In parallel, APCs and CD4+ cells also activate naïve CD8+ T cells which can migrate to the infection site and induce cytolysis of infected cells (Fig. 1D). Later, both CD4+ and CD8+ cells differentiate into memory T Cells (Fig. 1C, E). Affinity maturation for B and T cells takes several weeks [29–30], meaning that a typical primary infection is mostly resolved by the innate immune system (non-strain-specific cells and antibodies). However, any subsequent exposures to the same or antigenically similar antigens will result in a recall and boost of strain-specific memory cells leading to faster and better adaptive immune response [4,31].

**Figure 1:**
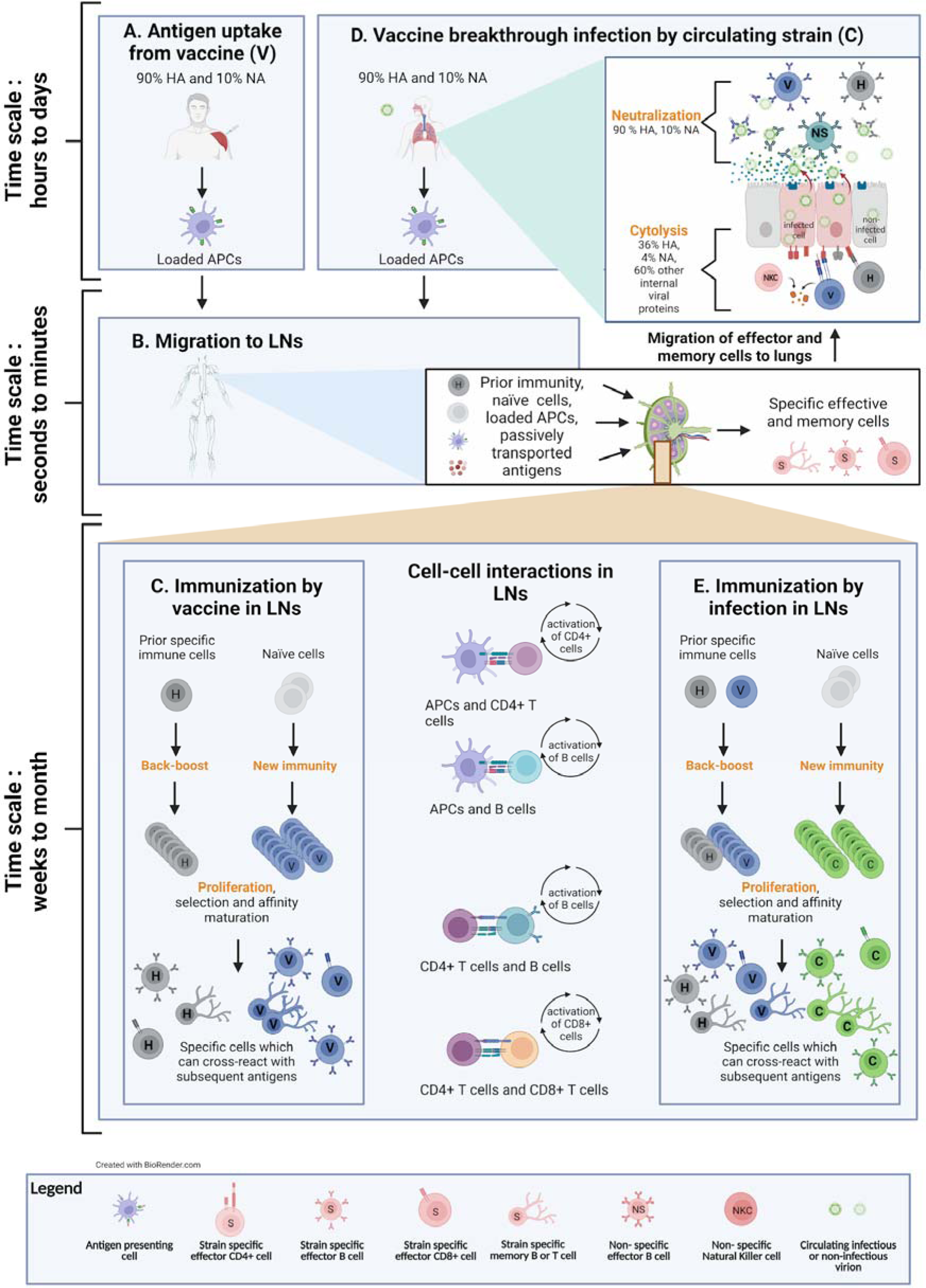
Overview of multi-strain model. **A.** Vaccine antigen uptake by antigen-presenting cells (APCs). **B.** Once loaded, APCs migrate from the injection site to the lymph nodes. Antigens (HA, NA) can also reach the lymph nodes by passive lymphatic drainage. **C.** Immunization in the lymph nodes results in a back-boost of prior immunity (historical strains, H) as well as the formation of new immunity specific to the vaccine strain (V). In the lymph nodes, several cell-cell interactions amplify the proliferation of specific immune cells, notably, the interaction between APCs and naïve B and CD4+ cells. Activated CD4+ cells also interact with B and CD8+ cells which differentiate into effector cells, with neutralizing and cytolytic functions respectively. **D.** Upon exposure to a seasonal circulating strain (C), specific immune cells migrate from the lymph nodes to the lungs. The different populations of immune cells interact with the new antigen C according to a cross-reactivity curve relating binding avidity constants to AgD between the new antigens and the old ones that elicited each strain-specific population. The rates of neutralization of specific antibodies elicited against H and V respectively depend on the AgD in HA and NA between H and C and V and C, weighted by the relative abundance of HA (90%) and NA (10%). 60% of pre-existing CD8+ cells have a rate of cytolysis which is independent of the AgD in HA and NA between previously encountered antigens (H, V) and C. If the specific immunity raised against H and V strains is sufficient to suppress the replication of the circulating strain or to control it without symptoms, the infection is respectively considered prevented or sub-clinical. **E.** In case of viral replication and appearance of symptoms corresponding to a vaccine breakthrough infection, there is an immunization against strain C, with a back-boost of the specific immunity against V and H. In case of severe infection lasting more than a few weeks, this new immunity against strain C can help resolve the infection.

Cross-reactivity in adaptive immune response was previously described by a model based on statistical mechanics [25], where the antigenic drift in the main epitopes of the viral surface proteins HA and NA resulted in a non-linear antibody response. This model was used to show that AgD between vaccine and circulating strains correlated well with VE against A/H3N2 from 1971 to 2004 [26]. It was estimated that the antibody affinity constant decreases non-linearly with AgD [27] and we assumed a similar relationship in our model. However, rather than using the number of epitope mutations, we define AgD as the output of the antigenic advance model described in Neher et al (2016) [32], normalized over the last 10 seasons (see Methods). This model, based on the relationship between strain genetic differences and their antigenicity, quantified by hemagglutination inhibition assays (HI titers), interprets antigenic data in a phylogenetic context [32]. Importantly, this model predicts the antigenic properties of strain pairs that have not been characterized experimentally [32]. Moreover, the AgD given by this model are well correlated to the number of epitope mutations [32].

To account for strains encountered by a virtual patient before or during a simulation, we consider strain-specific antibodies and strain-specific adaptive effector and memory cell populations co-existing in a patient (Fig. 1, Supplementary Fig. S1, Supplementary Table S1). The neutralization rate of cross-reactive specific antibodies and the proliferation of memory B cells depends strongly on AgD, whereas most of the cytolysis and proliferation of CD8+ cells does not, since a little more than half of the total CD8+ cells are targeted against internal viral proteins [33–34].

We simulate the pharmocokinetics of intramuscular injection of split vaccines by partitioning the initial HA and NA dose into direct lymphatic drainage and APC cell uptake [35]. The HD vaccine contains four times the antigens of the SD vaccine [36]. Given lack of data, our null hypothesis is that the number of primed APCs is linearly dependent on the dose.

To model infection severity, we consider the upper respiratory tract (URT) and the lower respiratory tract (LRT), the latter involving the airways below the larynx. We use a target cell limited within-host model [12–14,16,18], where the virus enters the body through the URT, infects lung epithelial cells, replicates inside them and can spread to the LRT. After activation, the immune system clears infected cells and viral particles located in the extracellular space (Fig. 1D). Outcomes at the patient level include infection, which can be symptomatic or not, with varying degrees of severity [37,38], and seroprotection [39], and are further defined in Table 1.

**Table 1.** Patient level outcome definitions used in the model.

|  | **Definition of outcomes** | **Reference** |
| --- | --- | --- |
| **Infection** | Patients are exposed at a variable time during the season to a single strain corresponding to the dominating strain of the main circulating clade of the season. However, each patient is exposed at the same time in the control and the treated (vaccinated) arm. The patient exposure dose is defined as the number of total viral particles per lung epithelial cells, also called multiplicity of infection. The dose to which a patient is exposed contains a variable percentage of infectious virions relative to non-infectious particles to mimic community transmission from individuals who are at variable stages of disease. If the maximum of total viral load exceeds the patient exposure dose, a patient is considered (subclinically or clinically) infected, as the viral replication exceeds its degradation by the immune system. | Jones et al (2020)<sup>38</sup> |
| **Seroprotection** | If the hemagglutinin inhibition (HI) titer is superior to 40 ($\log_2(\text{HI titer}) \geq 4$), the patient is considered seroprotected. This threshold has been historically considered as the 50% decreased risk in influenza infection. | Hobson et al (1972)<sup>40</sup> |
| **Presence/absence of symptoms** | As the range of nasal pro-inflammatory cytokines in symptomatic patients was reported as 2-130 pg/mL, in mild symptomatic young patients, we define the occurrence of symptoms as a simple threshold. If the pro-inflammatory cytokines in the URT remain below 2 pg/mL, the patient has no symptoms and the infection is considered asymptomatic (subclinical). Above this threshold, the infection is considered symptomatic (clinical). | Kaiser et al (2001)<sup>39</sup> |
| **URT/LRT infection** | If the viral concentration in the URT exceeds a calibrated threshold, the virus can spread to the LRT. The same threshold is used in infections by H1N1 and H3N2 for all patients. | Calibrated threshold |
| **Disease severity** | Symptomatic infections can be mild or severe. If the cumulative cellular damage exceeds a calibrated threshold in LRT infections, the infection is considered severe (requiring hospitalization), otherwise it is considered mild (only requiring a doctor visit). | Calibrated threshold |
| **Time to viral clearance** | Number of days necessary for the total infectious and non-infectious viruses to drop below a calibrated threshold in the URT and LRT. Used as a proxy for infection duration. | Calibrated threshold |

### Our model calibration process

We estimated the values of the parameters that could not be derived from literature via calibration with constraints defined by relevant data, based on *in vitro* and *in vivo* studies (Supplementary Fig. S2, see Methods). Using a covariance matrix adaptation evolution strategy, this step-by-step approach can be applied to a wide range of biological models [40]. We calibrated vaccine immunogenicity and viral dynamics independently. We then calibrated immune system dynamics in response to primary infection.

To calibrate pathogenesis in a population of non-naïve unvaccinated humans, we defined three reference patients exhibiting respectively an asymptomatic, mild symptomatic and severe symptomatic infection when exposed to a virus (Fig. 2). These reference patients, with typical duration of symptoms [41], were used to define the initial distributions of patient descriptors from which we simulated a large set of plausible patients (Supplementary Fig. S3).

**Figure 2:**
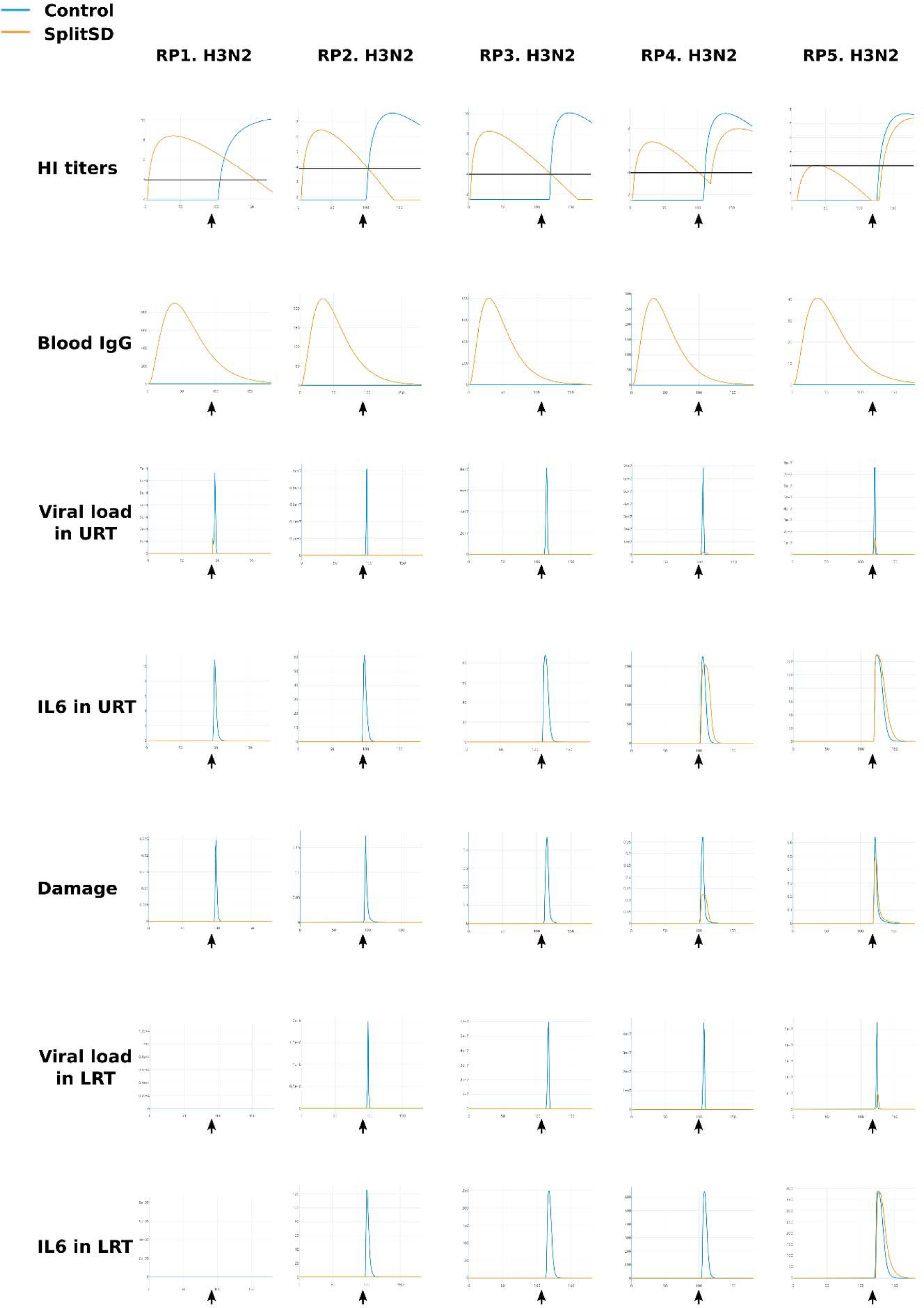
Calibration of reference patients. Time-courses of 7 main variables (rows) in 5 reference patients (RP) (columns) in control arm (blue) and split vaccine arm (orange, vaccination at day 1) with exposure to H3N2 at the indicated time points (arrows, between 80 and 120 days after vaccination) in both clinical arms over 180 days. All patients are aged between 70 and 80. First row: log2 HI titers raised against the vaccine strain. Second row: concentration of IgG specific to the vaccine strain in blood in nanomol/L. Third row: total viral load in the URT in mRNA/mL. Fourth row: concentration of pro-inflammatory cytokines (IL6) in the URT in nanomol/mL. Fifth row: instantaneous damage expressed as the fraction of infected lung epithelial cells compared to healthy lungs. Sixth row: total viral load in the LRT in mRNA/mL. Seventh row: concentration of pro-inflammatory cytokines (IL6) in the LRT in nanomol/mL. RP1: control asymptomatic infection and no vaccine breakthrough infection. RP2: control mild symptomatic infection and no vaccine breakthrough infection. RP3: control severe symptomatic infection and no vaccine breakthrough infection. RP4: control severe symptomatic infection and mild symptomatic vaccine breakthrough infection. RP5: control mild symptomatic infection and mild symptomatic vaccine breakthrough infection. All patients except RP5 reach seroprotective levels less than one month after vaccination (comparison of orange curve and black line in first row corresponding to log2 HI titers equals to 4). Vaccine breakthrough infections are identified as rebound of the HI titers after exposure in the vaccine arm (orange curve in RP4-5).

To simulate immunosenescence, we assumed a decrease in the number of naïve cells, an increase in pro-inflammatory cytokine autocatalysis (i.e interleukin 6) and a decrease in antiviral cytokine autocatalysis (i.e. interferon type III) with age [42–44]. The neutralization rate of antibodies was also assumed to decrease with age [45].

To calibrate the clinical effect of the SD vaccine at population level (Supplementary Fig. S3), we simulated a trial with a vaccine arm against a control arm without vaccine. A whole influenza season was simulated and patients were exposed, at a variable day, to A/H1N1 and A/H3N2 in two separate arms. We calibrated this four-arm trial using data from the 2010-2011 season, using the reported SD VE of 47 % (95% CI, 24%-63%) in 50+ adults [46]. The strains used in the 2010 vaccine were well matched to the circulating viruses [47] and the AgD between these and the circulating strains reported on Nextstrain [48] were small for HA and NA in both subtypes.

At the population level, we defined two primary clinical endpoints to allow comparison with randomized clinical trials (RCT) and real-world data (RWD, Table 2): the seroprotection rate [38] and the overall prevention of symptomatic infections, which is assumed to be comparable to VE quantified by surveillance centers in test-negative design studies using PCR-confirmed laboratory influenza virus infection. The comparison is only indicative since the definition of the control groups differs in RWD and in Virtual population (VP). In our virtual clinical trial, each patient is his own control, while in real data, control patients are tested negative to IAV but seek care at the same facilities as those who are tested positive.

**Table 2.**
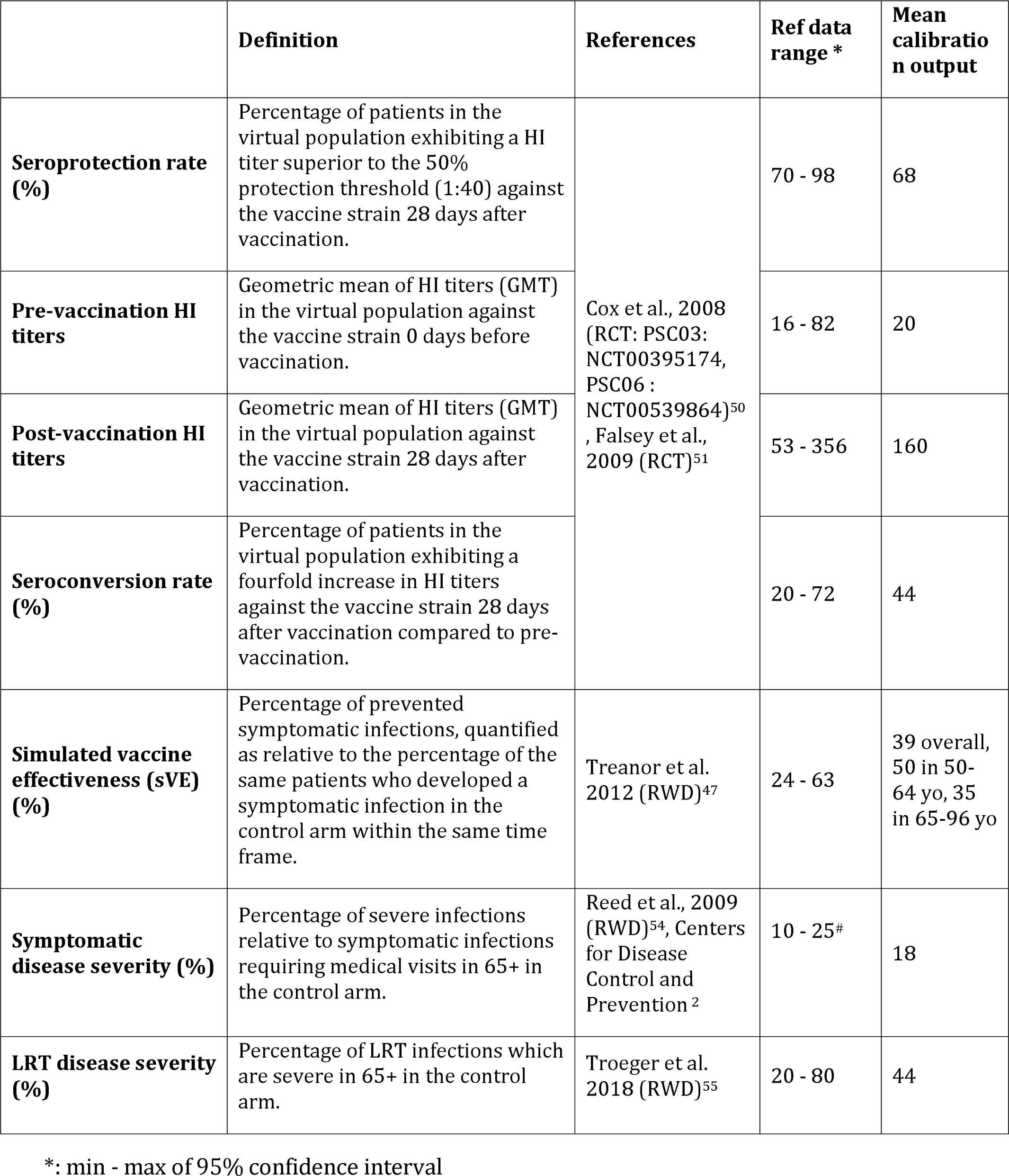
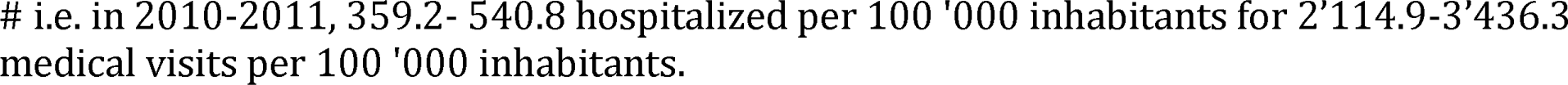
Population-level outcome definitions used in the model, ranges of data estimates reported in the literature and mean calibration output.

|  | **Definition** | **References** | **Ref data range \*** | **Mean calibration output** |
| --- | --- | --- | --- | --- |
| **Seroprotection rate (%)** | Percentage of patients in the virtual population exhibiting a HI titer superior to the 50% protection threshold (1:40) against the vaccine strain 28 days after vaccination. | Cox et al., 2008 (RCT: PSC03: NCT00395174, PSC06 : NCT00539864)<sup>50</sup>, Falsey et al., 2009 (RCT)<sup>51</sup> | 70 - 98 | 68 |
| **Pre-vaccination HI titers** | Geometric mean of HI titers (GMT) in the virtual population against the vaccine strain 0 days before vaccination. |  | 16 - 82 | 20 |
| **Post-vaccination HI titers** | Geometric mean of HI titers (GMT) in the virtual population against the vaccine strain 28 days after vaccination. |  | 53 - 356 | 160 |
| **Seroconversion rate (%)** | Percentage of patients in the virtual population exhibiting a fourfold increase in HI titers against the vaccine strain 28 days after vaccination compared to pre-vaccination. |  | 20 - 72 | 44 |
| **Simulated vaccine effectiveness (sVE) (%)** | Percentage of prevented symptomatic infections, quantified as relative to the percentage of the same patients who developed a symptomatic infection in the control arm within the same time frame. | Treanor et al. 2012 (RWD)<sup>47</sup> | 24 - 63 | 39 overall, 50 in 50-64 yo, 35 in 65-96 yo |
| **Symptomatic disease severity (%)** | Percentage of severe infections relative to symptomatic infections requiring medical visits in 65+ in the control arm. | Reed et al., 2009 (RWD)<sup>54</sup>, Centers for Disease Control and Prevention <sup>2</sup> | 10 - 25<sup>#</sup> | 18 |
| **LRT disease severity (%)** | Percentage of LRT infections which are severe in 65+ in the control arm. | Troeger et al. 2018 (RWD)<sup>55</sup> | 20 - 80 | 44 |

\*: min - max of 95% confidence interval

We used RCT immunogenicity data for SD vaccination in a 50-64 year old population [49] and in a 65+ years population [49–50], showing that at least 70% of patients in these age categories were seroprotected against A/H1N1 and A/H3N2 at 28 days post-vaccination (Table 2). By joining the parameter distributions of two reference patients - one succeeding and one failing to reach seroprotection levels post-vaccination (Fig. 2) - we refined the distributions of patient descriptors and generated a new set of plausible patients. Using published methods [51–52], we selected patients that allowed for the best reproduction of the reported distribution of seroprotection rate and simulated VE (sVE) against symptomatic infections in the SD arm against H1N1 and H3N2 infection in 50+ patients (Supplementary Fig. VpopCalibration). When sampling the most plausible patients, we aimed at generating a correlation between age and cumulative epithelial damage in the VP to simulate the higher rate of hospitalization in older adults reported by CDC. Rare or implausible patients who failed to clear the virus in the lungs within one month after infection in the control arm and in vaccine breakthrough cases were not sampled.

In the calibrated VP, sVE is higher in the 50-64 group than in the 65+ group against both subtypes (Table 2). The estimated geometric mean titers (GMT) against the vaccine strains pre-vaccination (t = 0) and post-vaccination (at 28 days) are within the range reported in RCTs (Table 2). The average seroconversion rate is 44% against both subtypes, which is in concordance with RCTs. The percentage of severe infections relative to symptomatic cases in the 65+ population is consistent with the data reported before [53]. The proportion of LRT infections which are severe and require hospitalization in 65+ are within the ranges reported worldwide [54].

### Simulations over consecutive seasons confirm the importance of vaccine match

#### Inputs

Systematic reviews and meta-analyses on efficacy and effectiveness of split vaccines in preventing influenza-associated clinical outcomes found that HD performs consistently better than SD vaccine in adults aged 65+ [55], but the relative VE (RVE) depended on whether seasons were dominated by A/H3N2 or A/H1N1 and on the antigenic match of the vaccine to the predominant circulating strains [8].

As the calibration used US estimations of the VE in 2010-2011 on 50+ adults, we performed seasonal simulations using the main circulating A subtype in the US between 2011 and 2021 reported by the CDC and the predominant clade that circulated mid-season in the USA as reported on Nextstrain [48] (Table 3). The input of each seasonal simulation is a set of normalized AgD in HA and NA between a sampled strain from the predominant clade and the corresponding seasonal vaccine strain.

**Table 3.**
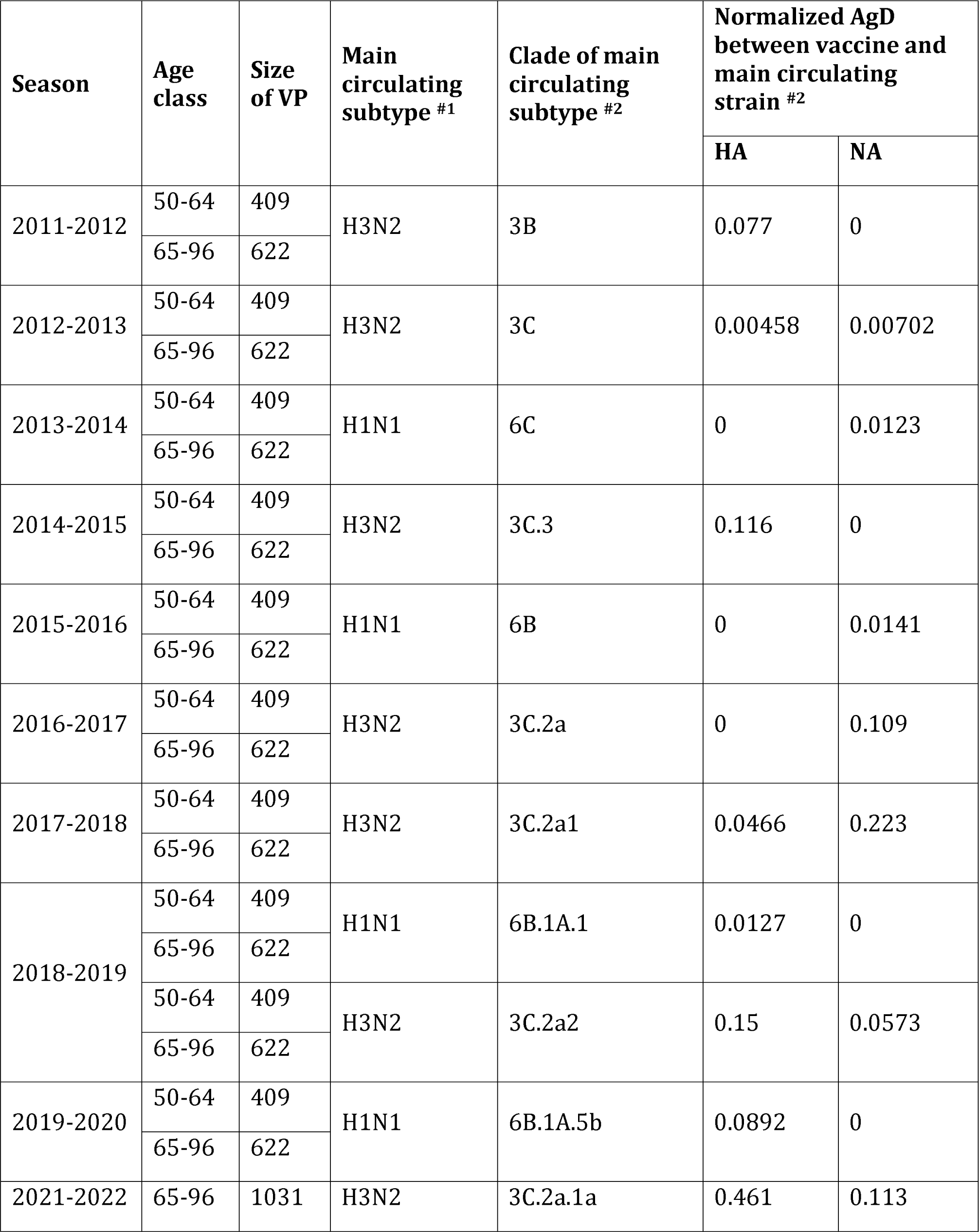
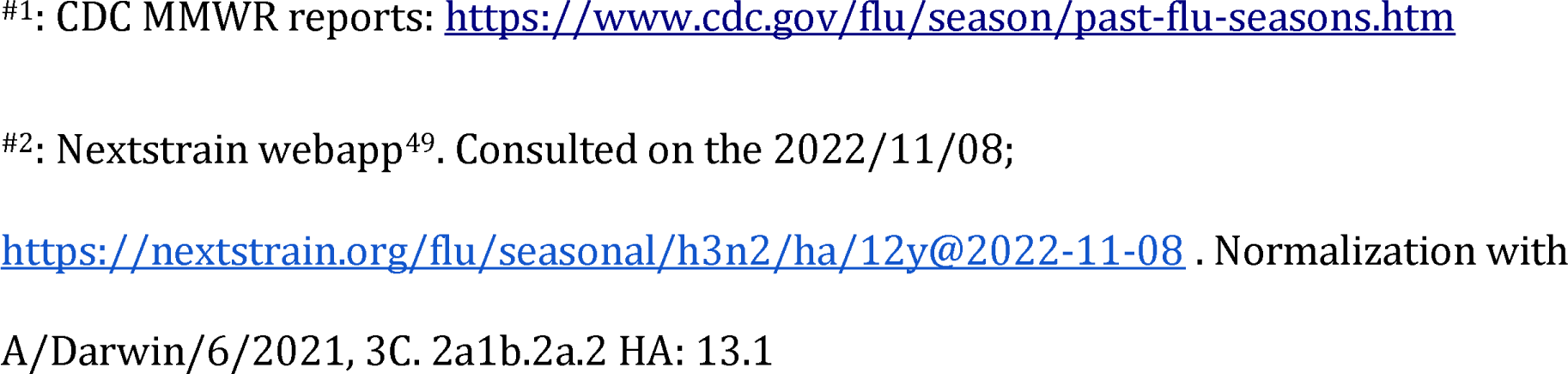
Model inputs used to simulate seasons, from 2011 to 2021, USA.

| Season | Age class | Size of VP | Main circulating subtype #1 | Clade of main circulating subtype #2 | Normalized AgD between vaccine and main circulating strain #2 |  |
| --- | --- | --- | --- | --- | --- | --- |
|  |  |  |  |  | HA | NA |
| 2011-2012 | 50-64 | 409 | H3N2 | 3B | 0.077 | 0 |
|  | 65-96 | 622 |  |  |  |  |
| 2012-2013 | 50-64 | 409 | H3N2 | 3C | 0.00458 | 0.00702 |
|  | 65-96 | 622 |  |  |  |  |
| 2013-2014 | 50-64 | 409 | H1N1 | 6C | 0 | 0.0123 |
|  | 65-96 | 622 |  |  |  |  |
| 2014-2015 | 50-64 | 409 | H3N2 | 3C.3 | 0.116 | 0 |
|  | 65-96 | 622 |  |  |  |  |
| 2015-2016 | 50-64 | 409 | H1N1 | 6B | 0 | 0.0141 |
|  | 65-96 | 622 |  |  |  |  |
| 2016-2017 | 50-64 | 409 | H3N2 | 3C.2a | 0 | 0.109 |
|  | 65-96 | 622 |  |  |  |  |
| 2017-2018 | 50-64 | 409 | H3N2 | 3C.2a1 | 0.0466 | 0.223 |
|  | 65-96 | 622 |  |  |  |  |
| 2018-2019 | 50-64 | 409 | H1N1 | 6B.1A.1 | 0.0127 | 0 |
|  | 65-96 | 622 |  |  |  |  |
|  | 50-64 | 409 | H3N2 | 3C.2a2 | 0.15 | 0.0573 |
|  | 65-96 | 622 |  |  |  |  |
| 2019-2020 | 50-64 | 409 | H1N1 | 6B.1A.5b | 0.0892 | 0 |
|  | 65-96 | 622 |  |  |  |  |
| 2021-2022 | 65-96 | 1031 | H3N2 | 3C.2a.1a | 0.461 | 0.113 |

#1: CDC MMWR reports: https://www.cdc.gov/flu/season/past-flu-seasons.htm
#2: Nextstrain webapp<sup>49</sup>. Consulted on the 2022/11/08;

Using the calibrated VP, we investigate how sVE depends on seasonal variation in AgD between the vaccine strain and the main circulating strain. Despite variable patient history, the VP is assumed to exhibit a constant level of prior immunity to the seasonal vaccine strain in every simulated season (See Methods). The model is also used to estimate how much RVE between the two doses depends on AgD in HA and NA when virtual populations from season to season are perfectly comparable in all their immune characteristics, including prior immunity.

#### Outputs

Our primary model outcome is sVE against symptomatic infections. Each patient, represented by a unique set of 60 descriptors, is simultaneously included in 3 different arms: control, splitSD and splitHD. This outcome is not directly comparable to VE based on RWD (Table 4). Indeed, the adjusted VE estimated from test-negative design studies on laboratory confirmed influenza cases (CDC Vaccine Effectiveness Studies) is usually stratified by viral subtype and by age, but not by vaccine type. Among US Medicare beneficiaries aged 65+, the proportion of individuals receiving HD increased considerably over the last decade [8]. Moreover, an increasing proportion of vaccinees received other vaccines (cell-based, recombinant or egg-based adjuvanted) in recent years [56]. Based on the market share of the vaccine manufacturer of split HD vaccine (Sanofi), we can estimate the percentage of vaccinees aged 65+ who received SD relative to HD [57] and derive a weighted sVE (wsVE) accounting for the percentage of vaccinees receiving SD each season. This improves the comparison with VE based on RWD but still does not account for all vaccine types.

**Table 4.**
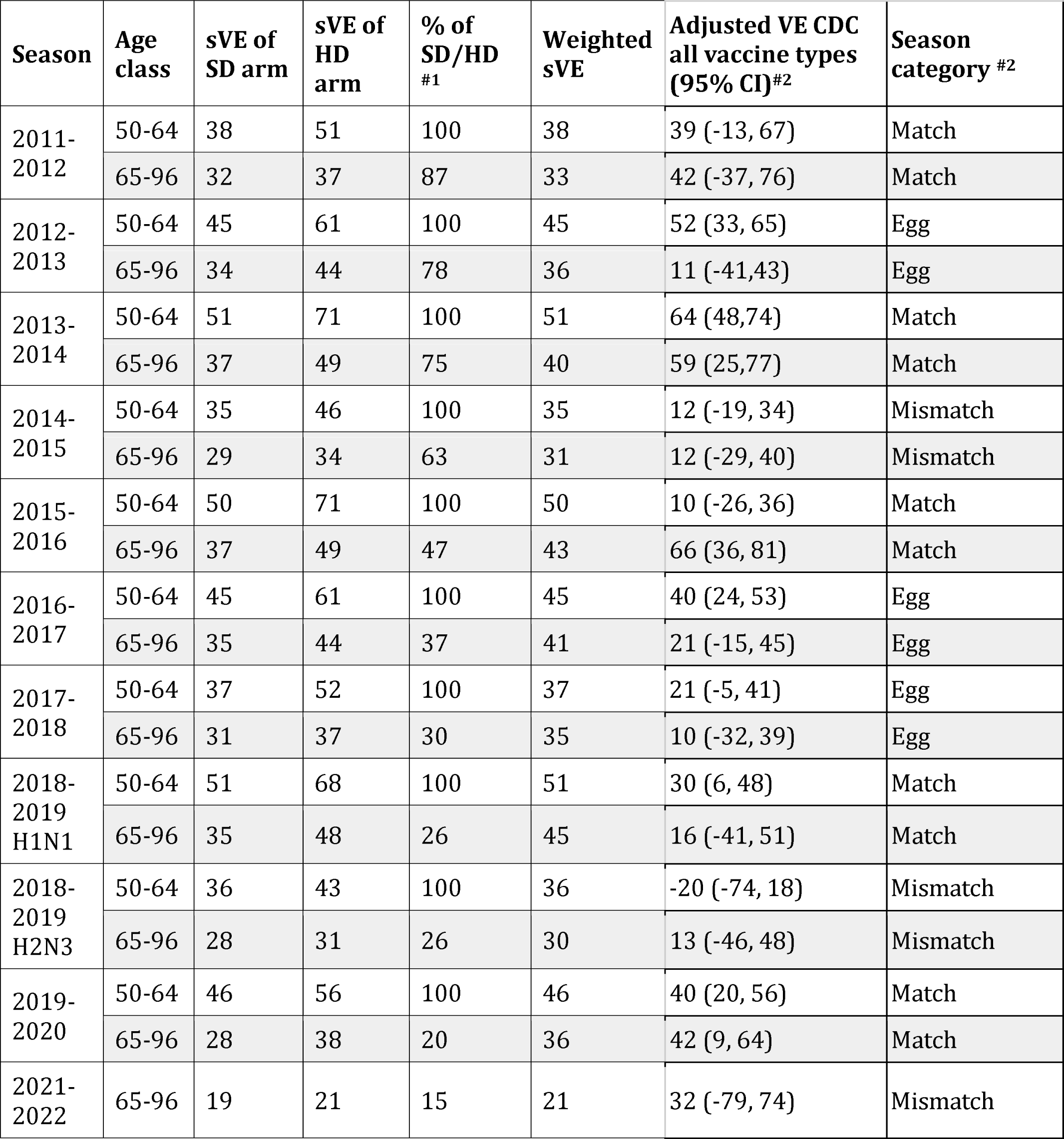
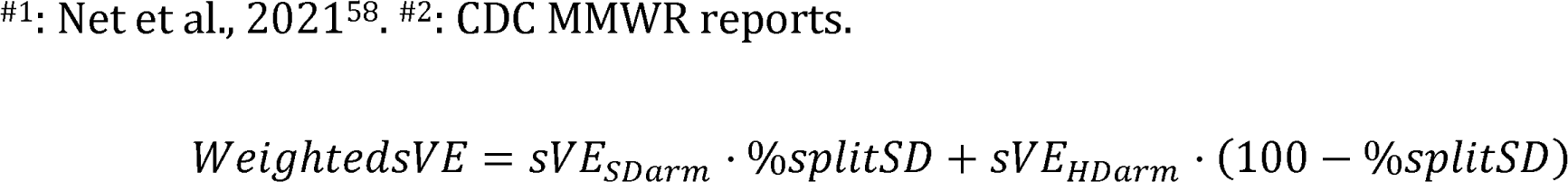
Outputs of simulated vaccine effectiveness (sVE) against symptomatic infections in vaccine arms and weighted sVE using the proportion of vaccinees receiving split standard dose (SD) rather than split high dose (HD) over consecutive seasons.

| Season | Age class | sVE of SD arm | sVE of HD arm | % of SD/HD #1 | Weighted sVE | Adjusted VE CDC all vaccine types (95% CI)#2 | Season category #2 |
| --- | --- | --- | --- | --- | --- | --- | --- |
| 2011-2012 | 50-64 | 38 | 51 | 100 | 38 | 39 (-13, 67) | Match |
|  | 65-96 | 32 | 37 | 87 | 33 | 42 (-37, 76) | Match |
| 2012-2013 | 50-64 | 45 | 61 | 100 | 45 | 52 (33, 65) | Egg |
|  | 65-96 | 34 | 44 | 78 | 36 | 11 (-41,43) | Egg |
| 2013-2014 | 50-64 | 51 | 71 | 100 | 51 | 64 (48,74) | Match |
|  | 65-96 | 37 | 49 | 75 | 40 | 59 (25,77) | Match |
| 2014-2015 | 50-64 | 35 | 46 | 100 | 35 | 12 (-19, 34) | Mismatch |
|  | 65-96 | 29 | 34 | 63 | 31 | 12 (-29, 40) | Mismatch |
| 2015-2016 | 50-64 | 50 | 71 | 100 | 50 | 10 (-26, 36) | Match |
|  | 65-96 | 37 | 49 | 47 | 43 | 66 (36, 81) | Match |
| 2016-2017 | 50-64 | 45 | 61 | 100 | 45 | 40 (24, 53) | Egg |
|  | 65-96 | 35 | 44 | 37 | 41 | 21 (-15, 45) | Egg |
| 2017-2018 | 50-64 | 37 | 52 | 100 | 37 | 21 (-5, 41) | Egg |
|  | 65-96 | 31 | 37 | 30 | 35 | 10 (-32, 39) | Egg |
| 2018-2019 H1N1 | 50-64 | 51 | 68 | 100 | 51 | 30 (6, 48) | Match |
|  | 65-96 | 35 | 48 | 26 | 45 | 16 (-41, 51) | Match |
| 2018-2019 H2N3 | 50-64 | 36 | 43 | 100 | 36 | -20 (-74, 18) | Mismatch |
|  | 65-96 | 28 | 31 | 26 | 30 | 13 (-46, 48) | Mismatch |
| 2019-2020 | 50-64 | 46 | 56 | 100 | 46 | 40 (20, 56) | Match |
|  | 65-96 | 28 | 38 | 20 | 36 | 42 (9, 64) | Match |
| 2021-2022 | 65-96 | 19 | 21 | 15 | 21 | 32 (-79, 74) | Mismatch |

#1: Net et al., 2021<sup>58</sup>, #2: CDC MMWR reports.

#### Sources of variation in vaccine effectiveness

VE for A/H3N2 is usually lower than that of A/H1N1[58]. This is also reproduced in our predictions, with A/H3N2 dominated seasons exhibiting the lowest sVE (Fig. 3A). This difference is mainly due to the most frequent vaccine mismatch in A/H3N2 dominated seasons used as inputs.

**Figure 3:**
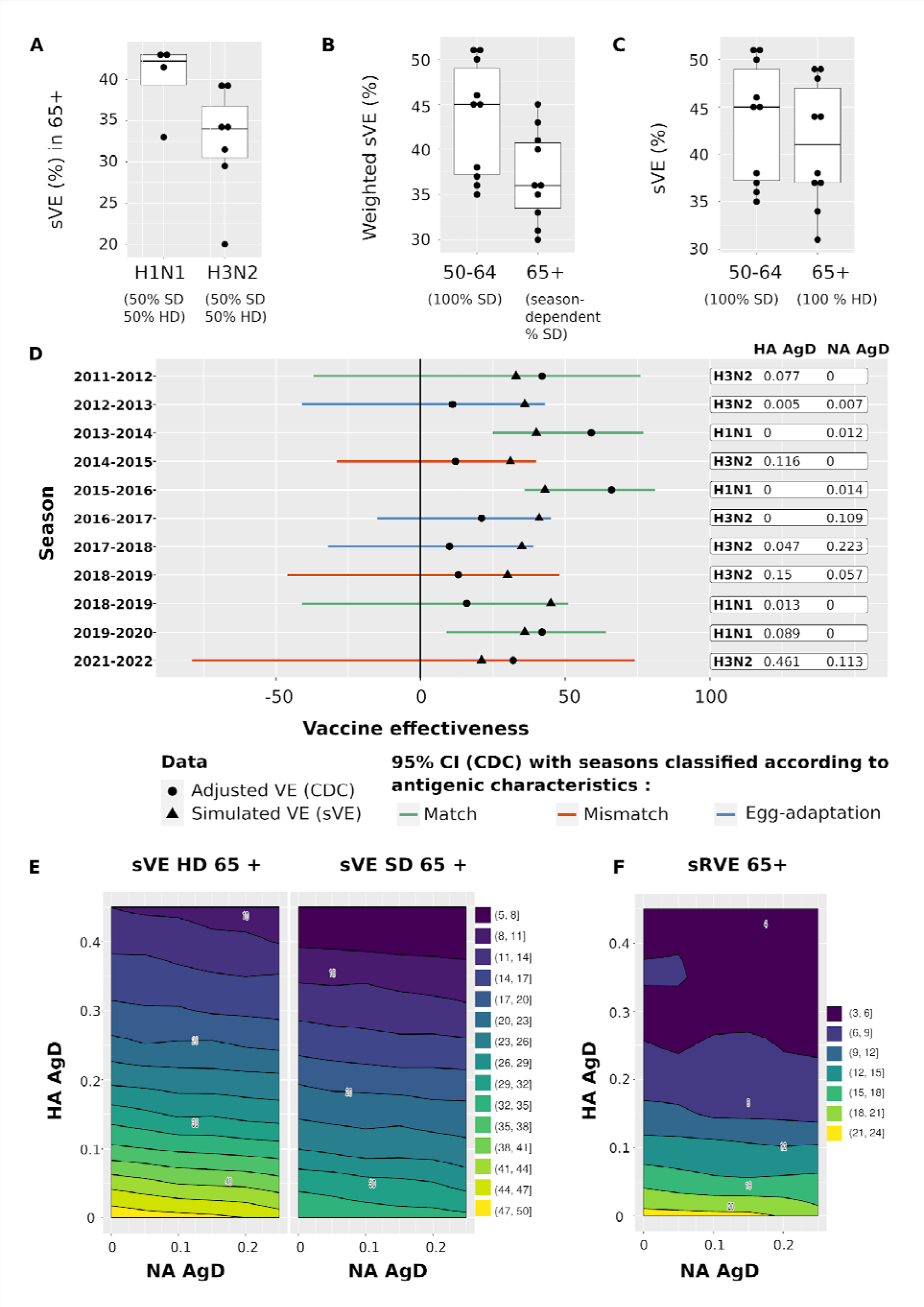
Model results. **A.** Comparison of sVE in 65+ in seasons dominated by A/H1N1 (4 seasons) and A/H3N2 (7 seasons) using pooled vaccine arms. Most seasons dominated by A/H3N2 exhibit a lower sVE than in A/H1N1 dominated seasons. **B.** Comparison of wsVE in younger and older age classes. 100% of patients aged 50-64 yo are vaccinated with splitSD as splitHD is not recommended in patients younger than 65 yo while the percentage of splitSD vaccinees relative to splitHD vaccinees in 65+ varies across seasons (Table 4). In those realistic conditions, the sVE in the 65+ is almost 10% lower than in the younger adults. **C.** If 100% of 65+ received the HD vaccine, the effect of immunosenescence would be almost canceled with respect to the younger adults receiving exclusively the SD vaccine. **D.** Timeline of wsVE against symptomatic infection (triangles) plotted over adjusted VE from CDC (dots) in 65+ between 2011 and 2022. The confidence intervals of adjusted VE are colored according to the antigenic characterization of main circulating seasonal strains with regards to the seasonal vaccine strains reported by the CDC as matched (green), mismatched (orange) and egg-adapted (blue) seasons. **E.** Heatmaps of predicted sVE against symptomatic infections as a function of AgD in HA and NA between the seasonal vaccine strain and the main seasonal vaccine strain, in 65+. The surface corresponds to theoretical seasons where combinations of AgD in HA and NA have been simulated to evenly sample the theoretical space of variation in antigenic distances observed over the last decade. The sVE of splitSD decreases with AgD in HA and NA, but the decrease is much faster with antigenic drift in HA than in NA. **F.** Although the sVE of splitHD is much higher than that of the splitSD, it decreases faster with antigenic drift, in particular in HA. **G.** The effectiveness of splitHD relative to splitSD (sRVE) decreases strongly with antigenic drift in HA and marginally in NA, but is nevertheless consistently different from 0, even at very large (and exceptional) combinations of AgD in HA and NA.

Figure 3B shows that the sVE against symptomatic infections, disregarding vaccine types, is lower in the 65+ than in the 50-64 group, illustrating the simulated immunosenescence. Our wsVE against symptomatic infections, considering only split vaccines, falls within CDC confidence intervals in all seasons in 65+. Most of the predicted wsVE in the 50-64 group also falls within these confidence intervals, except in seasons where VE is estimated to be equal or lower in this age group than in the 65+ group (2014-2015, 2015-2016, 2018-2019, Table 4). Indeed, 65+ patients most often exhibit the lowest VE [58], but not always, plausibly due to confounding factors of prior immunity. As we used exactly the same populations in all simulated seasons and calibrated our model to simulate a decreased sVE with age (Fig. 3B), failure is expected when the relationships between age and VE are inconsistent across seasons.

There is evidence suggesting that the level of protection of HD would be similar to that seen with SD in younger adults [59], which is also observed with our model (Table 4, Fig. 3C). Of note, sVE in HD arm has not been calibrated, and is the result of the assumed linear dose-response. Fig. 3D compares the simulated wsVE from 2011 to 2021 to the adjusted VE estimated from CDC.

To compare seasons, we use the classification between match, mismatch or egg-adaptation reported by CDC (Fig. 3D). Across seasons, our model predicts a smaller range of variation in sVE compared to adjusted VE (Fig. 3D). This is expected since our model assumes a constant level of prior immunity against each seasonal vaccine strain while prior immunity is an important confounding factor [4] hardly controlled for in RWD. As we disregarded egg-adaptation, the sVE for those seasons is overestimated. Disregarding these extra-sources of variation in VE, one can still see that the adjusted VE fluctuates from season to season, according to the vaccine match as described by CDC [8, 58] (Fig. 3D). The sVE over the whole combination of observed AgD in HA and NA follows this trend with a strong negative dependence of sVE on the input AgD in HA and a slighter dependence in NA (Fig. 3E).

#### Dependence of RVE on AgD

Importantly, the simulated RVE (sRVE) follows a similar decrease with AgD (Fig. 3E), ranging from 42% to 2.5% depending on season and age class. A double-blind RCT [59] reported a RVE against symptomatic infections of 24.2% (95% confidence interval [CI], 9.7 to 36.5) in 65+. It was noted also that the RVE estimates were higher in analyses restricted to cases caused by vaccine-similar strains, suggesting that RVE depends on AgD [59]. The sRVE quantified against hospitalization are listed for each season in Supplementary Table S3.

#### Improved immunogenicity with HD vaccine

The contribution analysis illustrates the patient descriptors correlated with a difference between the two vaccine arms, quantified with ’markers’ of response to vaccination (Fig. 4A-E). This sensitivity analysis is global, using the distributions and correlations of patient descriptors rather than varying each parameter independently [60]. Unsurprisingly, the dose-induced difference in seroprotection duration and blood IgGs is increased when antibody production rate is increased or antibody decay rate is decreased (Fig. 4A-B).

**Figure 4:**
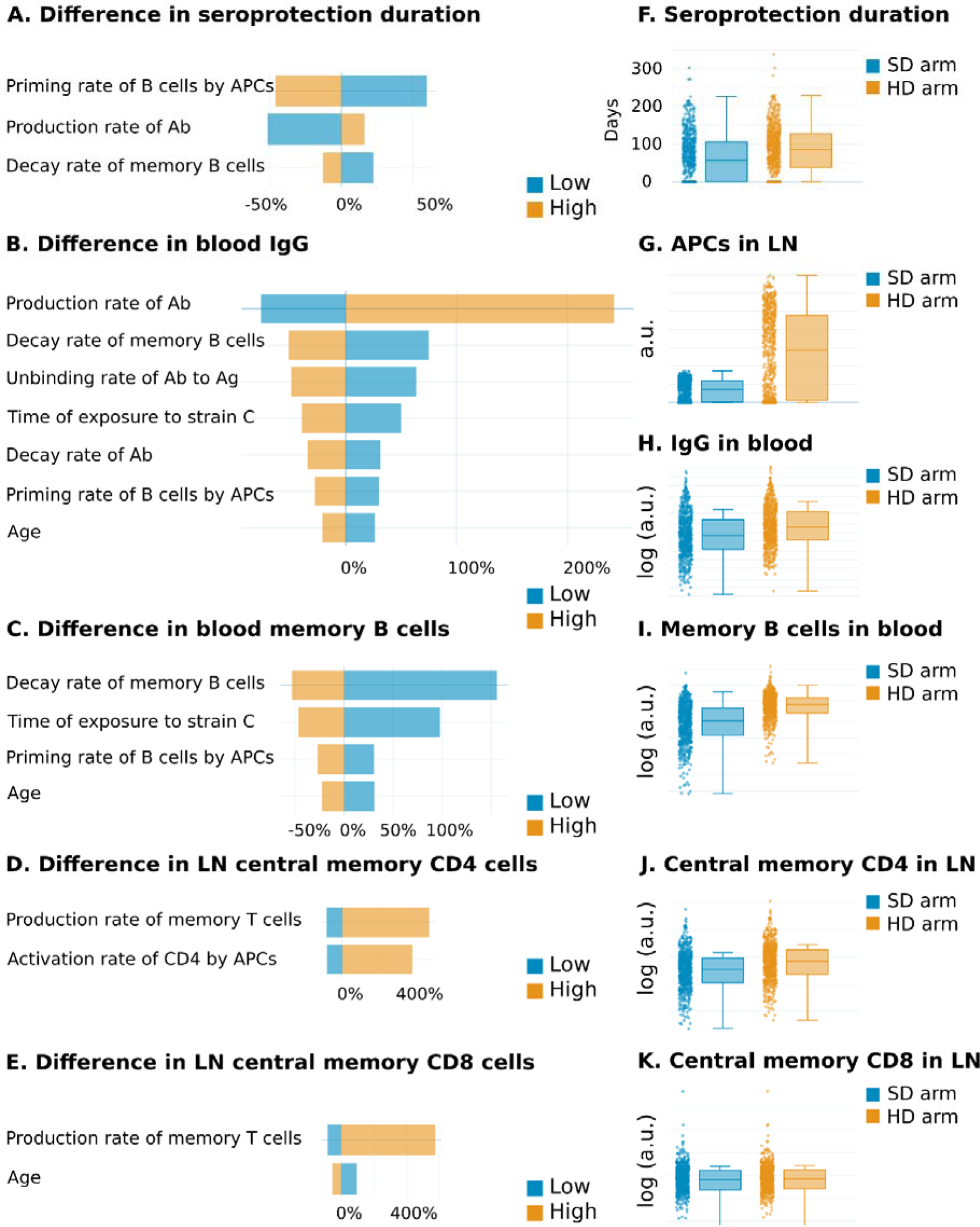
Predicted improvements with increased vaccine dose. **A-E.** Contribution analysis comparing, for each virtual patient, the difference between HD and SD arms, in seroprotection duration and vaccine-specific immunity at 28 days post-vaccination in 65+. A positive correlation between a marker of response to vaccination (i.e. dose-induced difference in seroprotection duration) is signified by low (blue) to high (orange) values from left to right, while a negative correlation goes from high to low values. Values are expressed in % change of the subpopulation’s median compared to the whole population’s median. For instance, in **A**, the subpopulation with the 50% highest values for the priming rate of B cells by APCs (“high” subpopulation) has a median for seroprotection duration 50% lower than the median of the overall population. For humoral immunity (**A-C**), the most sensitive parameters are related to the priming rate of B cells by APCs and to their antibody production and decay rates. While increasing the production rate of antibodies increases the differences between doses, increasing B cell priming and memory B cell decay rates decrease these differences. Increasing age (and thus immunosenescence) also decreases the difference between doses. Decreasing the time of exposure to viral antigens increases the differences between doses, due to back-boost of immunity against the vaccine strain, contributing to decreasing the difference between doses. For cellular immunity (**D-E**), the most sensitive parameters are related to the production rate of T cells which increases the difference between doses, while age decreases this difference. **F-K.** Quantified vaccine-specific markers tend to increase with vaccine dose, except central memory CD8 cells which show no change with vaccine dose. Distribution of vaccine-specific immunity in SD (blue) and HD (orange) arms in arbitrary units (a.u.), 28 days after vaccination. **F.** Seroprotection duration quantified as the number of days elapsed since vaccination where the HI titers remain superior to 1:40. **G.** APCs in lymph nodes. **H.** Antibodies specific to the vaccine strain in blood. **I.** Memory B cells in blood. **J.** Central memory CD4 cells in lymph nodes. **K.** Central memory CD8 cells in lymph nodes.

Unexpectedly, these differences are decreased when increasing age and the rate of B cell priming by APCs (Fig. 4C). Similarly, dose-induced differences in cellular immunity are increased when the production rate of T cells is increased and decreased with age (Fig. 4D-E). For antibody production rate, the tornado plot is asymmetrical, meaning that the relationship is non-linear: low antibody production rates result in 50% less dose-induced differences in seroprotection duration, while high rates result in 15% higher differences. Seroprotection duration is on average increased by 30% with HD vaccine compared to SD vaccine (Fig. 4F). This is attributable to the better priming of B and CD4 cells by APCs (Fig. 4G-J) which results in HI titers remaining above the 50% protection threshold for a longer time. As we assumed that the adaptive response partially depends on presentation of viral antigens by APCs to B and T cells, changes in parameters affecting APCs have a strong influence on seroprotection duration, which is consistent with a previous model [61].

## Discussion

Our model is highly sensitive to humoral and cellular immunity levels against historical strains, aligning with empirical and theoretical studies [4, 23, 31, 62]. In our seasonal simulations, we kept prior immunity constant, focusing on variations in the main circulating subtype and its antigenic distance (AgD) from the vaccine strain. This approach isolates the AgD’s impact on seasonal vaccine effectiveness (sVE). The sVE range may seem narrower than in real-world data (RWD) because we used the existing prediction of antibody cross-reactivity against an antigenic distance which is based on mutation counts and disregards phylogenetic relationships among strains [7, 25–27]. To refine our model, new data with recent strains (post-2010) is necessary for better calibration of cross-reactivity and newer definitions of AgD.

On one hand, our simulated relative vaccine effectiveness (sRVE) against hospitalization averages 43% for A/H1N1 and 46% for A/H3N2 (Supplementary Table S3), higher than RWD estimates of 5% to 30% depending on studies and seasons [56, 63–66]. On the other hand, our sRVE against symptomatic infections aligns with the lower range of randomized control trials (RCTs) [59]. Our method potentially deflates RVE estimates: using identical patients in different simulation arms avoids biases present in real-world clinical settings, like the at-risk vaccinee bias where high-dose (HD) vaccines are given to frailer adults [64–65]. Additionally, RVE against hospitalization can increase from negligible to 30% by precisely matching patients receiving standard-dose (SD) and HD vaccines by age and residence [64]. Our model’s predictions are in line with existing knowledge, validating it qualitatively as it accurately reflects immunosenescence, viral subtype, vaccine dose, and match effects.

Our model, however, does not account for egg-adaptation during vaccine production. In seasons where egg-adaptation was significant (2012, 2016, 2017 [9–11]), our model expectedly overestimates sVE based solely on AgD (Fig. 3D). Future iterations could differentiate the impact of strain selection and egg-adaptation on VE reduction.

Reducing seasonal influenza severity and preventing infection hinges on immune recognition of both HA and NA [7]. However, vaccination induces fewer anti-NA antibodies [67], and the quantity of NA in current vaccines is not standardized [68], with neuraminidase inhibition titers rarely measured. HA facilitates viral entry, while NA aids in viral release from cells [5]. Infection and vaccination result in varying ratios of anti-HA and anti-NA antibodies [69], but the underlying mechanisms of immunodominance are unclear. Therefore, our model assumes that the adaptive humoral response targets HA and NA based on their relative presence in infection and vaccination [69–70], overlooking other potential factors like immune cell hypermutation, selection, clonal expansion, or a difference in roles, not explicitly modeled here.

Our model does not consider cross-reactivity among subtypes, specifically the rare broadly neutralizing antibodies against the HA stalk [23, 67]. While childhood imprinting with a subtype reduces susceptibility to that subtype later, our model doesn’t account for age-based prior immunity. Consequently, it predicts lower VE against A/H3N2 than A/H1N1 (Fig. 3A) due to A/H3N2’s faster antigenic drift. Without considering anti-HA-stalk antibodies, the model is not equipped to predict age-based differential incidence by subtypes. Its application is confined to seasonal fluctuations over short time spans, not over a lifetime.

"Most evidence suggests that antibodies play a crucial role in the sterilizing immunity induced by vaccination, but T cell responses are also commonly stimulated [71]. Unlike humoral immunity, the cellular adaptive immune response primarily targets immunodominant epitopes in internal viral proteins, which are more conserved across A subtypes [34]. Therefore, long-term T cell immunity, especially from memory CD8+ cells, is likely to guard against reinfection by strains with different surface but identical internal proteins. In our model, we used the proportion of total T cell response attributable to internal versus surface viral proteins [33–34]. Consequently, antigenic distance (AgD) mainly impacts the humoral response, particularly to antigenic drift in HA, the primary antibody target (Fig. 3). However, the role of T cells in protection is poorly characterized, apart from CD4+ helper cells [71]."

Our model predicts that high-dose (HD) vaccines enhance seroprotection duration by more effectively priming APCs and activating CD4+ cells because of the linear dose-response relationship we assumed for APC priming (Fig. 4C). Only one study, an exploratory model in mice, examined APC priming’s dose-dependence [24]. It predicted a quasi-monotonic increase in seroprotection with higher IAV inactivated vaccine doses [24]. However, protection can also decrease as vaccine dose exceeds a certain threshold, leading to rapid antigen clearance by the innate immune system, preventing an effective adaptive response [24]. While our model’s predictions align with reported RVE over a decade using a linear dose-effect curve, further research is needed to clarify how different dosages impact APCs and T cells on a wider dose range.

The relationship between antibody titers and AgD has been investigated more thoroughly. For instance, it was demonstrated that the increase in antibody titers is greatest to the most recently encountered strain (as opposed to historical strains) but that antibody titers still spread over multiple antigenic clusters [4]. This broad subtype-specific back-boost and its relation to antigenic differences among strains was quantified in the form of antigenic landscapes [4]. Although the mechanism behind this back-boost is currently unknown, it appears more consistent with memory cell stimulation and antibody recall than a result of the production of novel antibodies with extensive cross-reactivity [4]. In our model, the back-boost is qualitatively consistent with this and other antibody landscape data [62]. By increasing the number of strains considered in this multi-strain model, it is now feasible to derive virtual quantitative antibody landscapes directly comparable to real patient-level antibody landscapes. Antibody landscapes thus appear as the most convenient high level descriptor of intra-population variation in humoral immunity both theoretically and in RCT/RWD. Despite substantial heterogeneity among the antibody landscapes of different individuals and highly variable individual response to vaccination, it was observed that each landscape shape was typically stable from one year to the next and had distinctive individual features [4, 62]. These observations suggest that most of the inter-patient variation in HI titers is due to variation in immune system and immunization history of patients. Our model further suggests that priming of APCs is important to account for inter-patient variability in HI titers. Moreover, it is often presumed that response to infection is broader or stronger than response to vaccination. Although a fair comparison of the antibody response to infection and vaccination is challenging [62], it seemed that the strength and breadth of the back-boost in response to infection and to vaccination were similar. Our model is compatible with such observations and could be pivotal in comparing further the mechanistic causes underlying the differential humoral responses to vaccination and infection in the future.

High-dose (HD) vaccines are licensed for people over 65 years to overcome pre-existing antibodies and immunosenescence. This enhanced vaccine has shown an increased capability of driving seroconversion and protection from influenza [72]. One hypothesis is that the increased antigen amount in HD vaccines prevents pre-existing antibodies from sequestering all antigens, enabling free antigens to activate memory B cells and thus promoting seroconversion against protective HA epitopes [59, 67].

In our model, a similar dose-effect mechanism is underlying the superiority of the HD vaccine in increasing seroprotective titers. However, this superiority depends on antigenic distance. If vaccine-induced antibodies poorly cross-react with the circulating strain due to a mismatch, the low cross-reacting antibodies might delay the production of better-matched antibodies. Particularly in seasons with significant antigenic mismatches in H3N2 dominant strains (2014, 2018, 2021), a small percentage (2-5%) of the virtual patients (VP) show a slightly longer viral clearance time than what is observed without vaccination. This aligns with the above hypothesis that binding of antigens by preexisting cross-reactive antibodies and memory cells sequesters antigens available for prime naïve B cells. While vaccine effectiveness is improved by increasing the match between vaccine and circulating strains, vaccine effectiveness is also boosted by reducing the match between vaccine strains and a patient’s pre-existing antibodies. In the context of SARS-CoV-2, it has been suggested that vaccine boosters using the beta-variant spike protein could provide better cross-neutralization against omicron variants than boosters based on recent omicron variant spike proteins, which built up herd immunity [73].

Our model confirms that HD consistently performs better than SD, against both subtypes, regardless of vaccine match, supporting the use of the HD vaccine in the older population. Research on antigen design concentrates on shifting natural immunodominance towards more broadly cross-reactive epitopes (i.e. headless antigens, HA-stalk) or prediction of the likely circulating strains based on pressure of selection. Besides the selection of vaccine strains, other active areas of research to improve effectiveness concentrate on vaccine designs which avoid egg-adaptation (i.e. cell-based and recombinant vaccines), but also glycosylation patterns (mRNA vaccines). Modeling and simulation is helping research in all these aspects, as well as supporting the choice of the optimal antigen dosage, especially in the susceptible populations.

## Methods

### Multi-strain model description

The multi-strain model is described by a system of ordinary differential equations (ODEs) and uses a virtual population approach where parameters are described by statistical distributions rather than scalar values, in order to represent different sources of variability [74]. Each virtual patient corresponds to a vector of parameter values drawn from the corresponding statistical distribution.

The multi-strain model is based on 4 independent submodels, which can run independently or in combination (Supplementary Fig. S1; Supplementary Methods):

1. The Immunization submodel describes building of a fast innate response and a slow adaptive response in lymph nodes and blood after antigen encounter with a time scale of days and weeks respectively (Fig. 1C; Fig. 1E),

2. The Vaccine Immunogenicity submodel describes the vaccine antigen-uptake by APCs at injection site (muscle) with a time-scale of hours to days (Fig. 1A),

3. The Viral Life Cycle describes the within-host infection and replication in lung epithelial cells with a time scale of hours to days (Fig. 1D) and

4. The Pathogenesis model describes the effector response to viral exposure (neutralization, cytolysis, inflammation) in the URT and LRT with a time scale of hours to days (Fig. 1D).

Once connected, these submodels allow the simulation of a variety of scenarios and conditions (Supplementary Fig. S1). The assumptions of these submodels are presented in Supplementary Methods.

In the multi-strain model, the different populations of strain-specific immune cells, antibodies and antigens have been multiplied according to the number of strains considered in a simulation (Fig. 1). Here, we consider 3 strains: 1) one historical strain (H) corresponding to a former circulating strain that a patient encountered 5 to 10 years before the start of the simulation (with remaining specific memory cells) ; 2) one vaccine strain (V) corresponding to a vaccine administered at the start of the simulation ; 3) one seasonal circulating strain (C) which can be encountered at a different time for each patient. This strain is used to test the vaccine effectiveness at the population level at the end of the season.

#### Cross-reactions

The intensity of cross-reactions between a strain and adaptive immunity elicited against another strain depends on the antigenic distance among strains. Antigenic distances between two strains are defined according to the Nextstrain platform [48] - a web browser-based application - that visualizes antigenic data on a continuously updated phylogeny, allowing to make their model outputs readily available. We used the antigenic distances between our reference virus of 2009 (A/H1N1/California/2009 and A/H3N2/Perth/2009) and the vaccine strains used in a particular season (after 2009), using the so-called antigenic advance submodel. We normalized all antigenic distances for HA and NA in A/H3N2 and A/H1N1 using the antigenic distance between A/H3N2/Wisconsin/67/2005 and A/H3N2/Darwin/6/2021 (clade 3C. 2a1b.2a.2).

The relationship between antigenic distance and antibody avidity constant is derived from Deem and Lee (2003) [25]. For simplicity, we simulate only the cross-reactivity at the main epitope of HA and NA proteins, as those authors assumed previously [7,25–27]. To describe the exponential decrease in antibody avidity constant with normalized antigenic distance between strain pairs and to infer the cross-avidity constant of strain-specific antibodies against other strains antigens, we use the following equation:

if HAagD <= 0.6

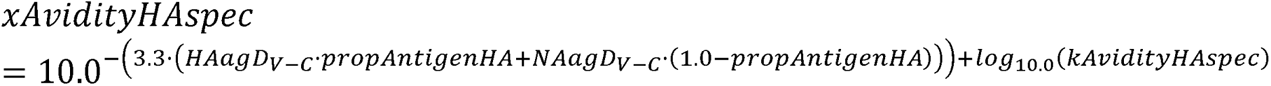

else

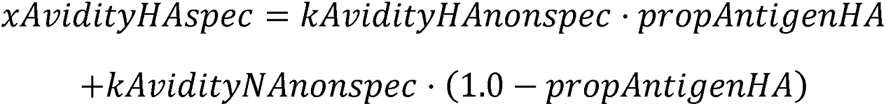

If the antigenic distance in HA main epitope is below 0.6 [25], the antibodies can cross-react with those antigens. Above this threshold, the antibodies do not cross-react with antigens and are assumed to have the same avidity as non-specific antibodies. In our model, the neutralization rate of cross-reactive specific antibodies as well as the proliferation of memory B cells depend strongly on antigenic distance in accordance with the relative abundance of HA and NA (where propAntigenHA equals 0.9) in a virion or split vaccine [69–70]. The cytolysis rate of CD8+ cells and their proliferation depend less strongly on antigenic distance, since only 40 % of the total CD8+ cells are targeted against viral surface protein epitope [33–34]. We thus assume that 36% of CD8+ cells response is targeted against HA, while 4% is targeted against NA (0.9*0.4 and 0.1*0.4 respectively).

The multi-strain model disregards non-neutralizing antibodies as well as antibodies against HA-stalk (Supplementary Methods). It thus considers only the antibodies raised against the main epitopes of HA-head and NA, using the non-linear relationship between the cross-avidity constants and normalized antigenic distance (the cross-reactivity equation for NA is the same as for HA because specific and non-specific avidities are assumed to be the same regardless of the antigen, see Supplementary Table S2). This model is phenomenological in aspects that relate to the relationship between the affinity of T cell receptor (TCR) and the effector functions of T cells (cytolysis, helper function). Despite extensive experimental work on TCR affinity, we were not able to establish a clear correlation between affinity and T-cell response because the available data are far from conclusive and even contradictory [75] (Supplementary Methods).

#### Simulation of hemagglutination inhibition (HI) assay

In clinical trials, HI assays are usually performed 28 days post-vaccination, as a correlate of protection. This assay quantifies a combination of quantity (concentration) and quality (avidity) of neutralizing antibodies developed in response to vaccination. HI titer refers to the highest serum dilution that fully inhibits hemagglutination due to antibody binding [76]. We used the theoretical model of Linnik et al (2022) [76] to predict the log2 HI titers from values of the concentration of specific neutralizing antibodies in serum (`Blood.ig`) and their avidity for HA (`kAvidityHAspec`) by fitting their phase diagram [76] :

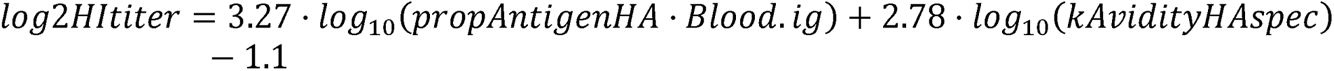

where `propAntigenHA` represents the relative proportion of HA antigens with respect to NA antigens in natural infection or in vaccination [60–70] (0.9). This equation returns continuous non-integer values of log2 HI titers to be compared to integer values returned by real HI assays. This equation is valid when the patient serum contains only one population of specific antibodies which were generated upon one antigen encounter, like in naïve patients.

To derive the log2 HI titers in patients who had successive immunizations, we compute a cross-reactive log2 HI titer. Each population of specific antibodies and new antigens are tested together using cross-reactivity and antigenic distances among past and new antigens. The cross-reactive HI titers against the historical (H), vaccine (V) and circulating strains (C) are simulated as the maximum of the HI titers simulated against each strain individually. We use the same equation as the HI titers described above, except that the cross-reactive equation `xLog2HItiter` uses the pairwise cross-avidity of antibodies for HA antigens rather than their avidity for HA.

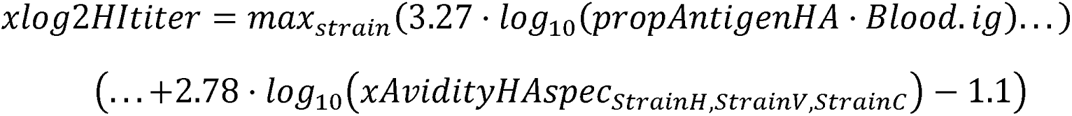

#### Initialization of a prior immunity with the historical strain

An individual’s previous antigen exposure through vaccination or infection may lead to a baseline level of immunity against influenza which may be highly heterogeneous across a population [77]. The initialized baseline level is assumed to be specific to a generic historical (H) strain that represents remaining immunity against both H1N1 and H3N2 viral subtypes (possibly from several strains within these subtypes). Strain H specific variables which are initiated (non-null) at the beginning of the simulation are: specific antibodies in blood, memory B cells in lymph nodes and blood, memory T cells in lymph nodes and tissue-resident memory T cells in the upper/lower respiratory tract. Because we calibrated the Influenza Viral Life Cycle Submodel using experimental data on the circulating A/H1N1/7/California/2009 and A/H3N2/Perth/2009 (Supplementary Methods), these strains are the oldest ones that can be encountered by a patient in our simulations. So here, we define prior immunity as immunization generated by strains encountered before 2009.

### Multi-strain model calibration

Calibration is an automatic numerical procedure, in which *a priori* unknown parameter values are concomitantly estimated and refined to reproduce desired model behavior, usually via an iterative process. Calibration constrains the dynamic behavior of the model by finding a set of parameter values that allows the model to represent biological behaviors consistent with literature [40]. All raw data used in the study was extracted from scientific publications and public CDC reports, no administrative permission was required for the access. We calibrate each submodel independently using data described in Supplementary Methods (steps 1 to 4). The integrated multi-strain model is calibrated to derive 3 reference patients exhibiting a spectrum of disease severity without treatment [41] and 2 reference patients exhibiting vaccine breakthrough infections with and without achieving seroprotective titers 28 days after splitSD vaccination (Fig. 2, Supplementary Methods, step 5). These 5 reference patients are then used to define plausible distributions of patient descriptors (Supplementary Fig. S2). Following the select and sample method [51–52], the virtual population is refined in an iterative process to match the immunosenescence, seroprotection rate, and proportions of prevented symptomatic infections of splitSD described in Supplementary Methods (steps 6 to 8) and Supplementary Fig S2.

### Multi-strain model simulations over consecutive seasons

The simulations are run on Jinko.ai, Novadiscovery’s proprietary platform, which uses the Sundials library [78], using LLVM evaluator and BDF solver, and relative and absolute tolerances of 0.000001. Numerical solutions of the system of ODEs as a function of time, as well as all remaining calculations and plots are performed using Jinko (Novadiscovery). The system of ODEs for the multi-strain model is available from the authors upon reasonable request.

In all seasons, we use the same VP in control and two vaccine arms. Thus, patients have the same prior immunity to the vaccine strain at the beginning of each season. From season to season, solely the encountered viral subtype and its antigenic distance with the vaccine strain changes, as reported by the CDC MMWR reports and antigenic characterization of viruses which circulated each season (Table 3).

We output the estimated HI titers against the historical and vaccine strains at the beginning of simulation (pre-vaccination, t = 0) and post-vaccination (at 28 days after the beginning of simulation). Our primary clinical outcome is the proportion of prevented symptomatic infections (mild and severe infections are pooled, Table 2). Our secondary clinical outcome is the proportion of prevented severe symptomatic infections only. In our model, severe symptomatic infections, mostly involving the LRT, are used as a proxy for hospitalization [54]. The proportion of prevented severe symptomatic infections is quantified as the percentage of severe symptomatic infections in vaccine arms, relative to the percentage of patients with severe symptomatic infections in the control arm.

The relative vaccine effectiveness (sRVE) is defined [79] as:

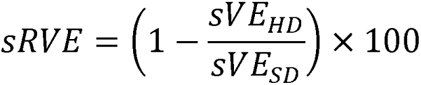

where sVE is calculated against all symptomatic infections or severe symptomatic infections only, using the number of prevented events in treated arms relative to the control arm within the same time window.

### Multi-strain model analysis in Jinko

#### Analysis

The visualization of time-series, boxplots and histograms comparing trials arms as well as the contribution analyses are done on Jinko.

Times series show the evolution of the selected clinical output(s) on a given time period, which is the trial duration selected during the configuration of your trial (Fig. 2).

##### Contour plots of sVE and sRVE

The surface of illustrated contour plots (Fig. 3) corresponds to theoretical seasons where combinations of AgD in HA and NA have been simulated to evenly sample (every 0.025 increment) the theoretical space of variation in antigenic distances observed over the last decade. These plots were generated with R from the data downloaded from Jinko.

Boxplots and Histograms (Fig. 4) give a representation of *scalar results* among the population. *Scalar Results* are the result of a reduce measure applied on clinical outputs. Most frequent measures used in Jinko are the Value of the output at a given time point, the minimum, the maximum, the area under the curve or the average of the output over the simulation period.

##### Contribution analysis

Contribution analysis (Fig. 4) is based on the comparison of statistical properties of subgroups of the VP versus properties of the whole VP using a quantity of interest (QOI). To compute one contribution analysis for a given QOI, the following process is applied:

We first compute the median of the QOI among the patients for each input descriptor.

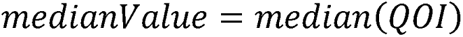

Then patients are sorted by increasing order of input descriptor value, and the population is split into two groups for which we compute the *lowMedianValue* and *highMedianValue* of the QOI. The relative contribution of the descriptor in the two groups is defined as :

In group 1, we have:

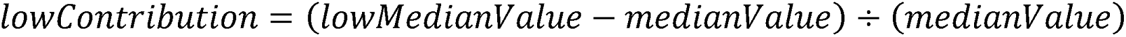

In group 2, we have:

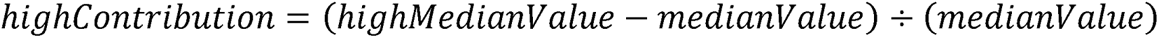

We center the Tornado graph on *medianValue* and the bars around corresponds to *lowContribution* and *highContribution*.

## Contributions

L.B., S.U., L.C., J-P.B., and E.C. supervised the study. S.U., M.H., A.I.T., N.R., E.J., E.P., J-B.G., E. C. & L.B. developed the model, performed simulations, and analyzed the results. S.U. and L.B. wrote the manuscript. S.S.C, L.C., E.T. and J-P.B critically revised the manuscript for important intellectual content. All authors contributed to the discussion of the results, reviewed the manuscript and gave final approval of the version to be published.

## Acknowledgements

The authors would like to thank the rest of Novadiscovery biomodeling team for their advice and feedback on the manuscript and scientific content, and the Scientific Software Engineering team for their work on the Jinko platform and other tools used.

The authors also acknowledge scientific contributions to the model from members of Sanofi team.

## Data availability

The dataset generated and analyzed during the current study, the system of ODEs for the multi-strain model and each submodel in SBML are available from the corresponding author upon reasonable request.

## References

1. Macias, A. E. et al. The disease burden of influenza beyond respiratory illness. Vaccine 39, A6–A14 (2021).

2. Centers for disease control and prevention. Disease burden of flu (2022), available from: https://www.cdc.gov/flu/about/burden/index.html. Accessed: 5^th^ of May 2023.

3. Smith, D. J. et al. Mapping the antigenic and genetic evolution of influenza virus. Science 305, 371–376 (2004).

4. Fonville, J. M. et al. Antibody landscapes after influenza virus infection or vaccination. Science 346(6212), 996–1000 (2014).

5. Krammer, F. The human antibody response to influenza A virus infection and vaccination. Nature Reviews Immunology 19(6), 383–397 (2019).

6. Hensley, S. E. et al. Hemagglutinin receptor binding avidity drives influenza A virus antigenic drift. Science 326(5953), 734–736 (2009).

7. Muñoz, E. T. & Deem, M. W. Epitope analysis for influenza vaccine design. Vaccine 23(9), 1144–1148 (2004).

8. Lee, J. K. H., Lam, G. K. L., Shin, T., Samson, S. I., Greenberg, D. P. & Chit, A. Efficacy and effectiveness of high-dose influenza vaccine in older adults by circulating strain and antigenic match: An updated systematic review and meta-analysis. Vaccine 39, A24–A35 (2021).

9. Skowronski, D. M., et al. Low 2012–13 influenza vaccine effectiveness associated with mutation in the egg-adapted H3N2 vaccine strain not antigenic drift in circulating viruses. PLoS ONE 9(3), e92153 (2014).

10. Zost, S. J. et al. Contemporary H3N2 influenza viruses have a glycosylation site that alters binding of antibodies elicited by egg-adapted vaccine strains. Proceedings of the National Academy of Sciences 114(47), 12578–12583(2017).

11. Liu, F. et al. Age-specific effects of vaccine egg adaptation and immune priming on A(H3N2) antibody responses following influenza vaccination. Journal of Clinical Investigation 131(8), (2021).

12. Baccam, P., Beauchemin, C., Macken, C. A., Hayden, F. G. & Perelson, A. S. Kinetics of influenza A virus infection in humans. Journal of Virology (2006).

13. Saenz, R. A. et al. Dynamics of influenza virus infection and pathology. Journal of Virology 80(15), 7590–7599 (2010).

14. Pawelek, K. A., Huynh, G. T., Quinlivan, M., Cullinane, A., Rong, L. & Perelson, A. S. Modeling within-host dynamics of influenza virus infection including immune responses. PLoS Computational Biology 8(6), e1002588 (2012).

15. Lukens, S. et al. A large-scale immuno-epidemiological simulation of influenza A epidemics. BMC Public Health 14, 1–15 (2014).

16. Simon, P. F. et al. Avian influenza viruses that cause highly virulent infections in humans exhibit distinct replicative properties in contrast to human H1N1 viruses. Scientific Reports 6(1), 24154 (2016).

17. Zarnitsyna, V. I., Handel, A., McMaster, S. R., Hayward, S. L., Kohlmeier, J. E. & Antia, R. Mathematical model reveals the role of memory CD8 T cell populations in recall responses to influenza. Frontiers in Immunology 7, 165 (2016).

18. Yan, A. W. C., Zhou, J., Beauchemin, C. A. A., Russell, C. A., Barclay, W. S. & Riley, S. Quantifying mechanistic traits of influenza viral dynamics using *in vitro* data. Epidemics 33, 100406 (2020).

19. Chen, X., Hickling, T. & Vicini, P. A mechanistic, multiscale mathematical model of immunogenicity for therapeutic proteins: Part 1 - theoretical model. CPT: Pharmacometrics & Systems Pharmacology 3(9), 1–9 (2014).

20. Giorgi, M., Desikan, R., Graaf, P. H. & Kierzek, A. M. Application of quantitative systems pharmacology to guide the optimal dosing of COVID-19 vaccines. CPT: Pharmacometrics & Systems Pharmacology 10(10), 1130–1133 (2021).

21 Desikan R., Linderman, S. L., Davis, C., Zarnitsyna, V. I., Ahmed, H. & Antia, R. Vaccine models pre,dict rules for updating vaccines against evolving pathogens such as SARS-CoV-2 and influenza in the context of pre-existing immunity. Frontiers in Immunology 13, 985478 (2022).

22. Alexandre, M. et al. Modelling the response to vaccine in non-human primates to define SARS-CoV-2 mechanistic correlates of protection. eLife 11, e75427(2022).

23. Zarnitsyna, V. I., Lavine, J., Ellebedy, A., Ahmed, R. & Antia, R. Multi-epitope models explain how pre-existing antibodies affect the generation of broadly protective responses to influenza. PLOS Pathogens 12(6), e1005692 (2016).

24. Handel, A., Li Y., McKay, B., Pawelek, K. A., Zarnitsyna, V. & Antia, R. Exploring the impact of inoculum dose on host immunity and morbidity to inform model-based vaccine design. PLOS Computational Biology 14(10), e1006505 (2018).

25. Deem, M. W. & Lee, H. Y. Sequence space localization in the immune system response to vaccination and disease. Physical Review Letters 91(6), 068101 (2003).

26. Gupta. V., Earl. D. J. & Deem. M. W. Quantifying influenza vaccine efficacy and antigenic distance. Vaccine 24(18), 3881–3888 (2006).

27. Deem. M. W. & Hejazi. P. Theoretical aspects of immunity. Annual Review of Chemical and Biomolecular Engineering 1, 247–276 (2010).

28. Mueller. S. N., Gebhardt. T., Carbone. F. R. & Heath. W. R. Memory T cell subsets, migration patterns, and tissue residence. Annual Review of Immunology 31, 137–161 (2012).

29. Yewdell. W. T., et al. Temporal dynamics of persistent germinal centers and memory B cell differentiation following respiratory virus infection. Cell Reports 37(6) (2021).

30. Woodland. D. L. & Kohlmeier. J. E. Migration, maintenance and recall of memory T cells in peripheral tissues. Nature Reviews Immunology 9(3), 153–161 (2009).

31. Yan. A. W. C., et al. Modelling cross-reactivity and memory in the cellular adaptive immune response to influenza infection in the host. Journal of Theoretical Biology 413, 34–49 (2016).

32. Neher. R. A., Bedford. T., Daniels. R. S., Russell. C. A. & Shraiman. B. I. Prediction, dynamics, and visualization of antigenic phenotypes of seasonal influenza viruses. Proceedings of the National Academy of Sciences 113(12), E1701–E1709 (2016).

33. Hayward. A. C., et al. Natural T cell–mediated protection against seasonal and pandemic influenza. Results of the flu watch cohort study. American Journal of Respiratory and Critical Care Medicine 191(12), 1422–1431 (2015).

34. Reber. A. J., Music. N., Kim. J. H., Gansebom. S., Chen. J. & York. I. Extensive T cell cross-reactivity between diverse seasonal influenza strains in the ferret model. Scientific Reports 8(1), 6112 (2018).

35. Moyer. T. J., Zmolek. A. C. & Irvine. D. J. Beyond antigens and adjuvants: Formulating future vaccines. Journal of Clinical Investigation 126(3), 799–808 (2016).

36. Gravenstein. S., et al. Comparative effectiveness of high-dose versus standard-dose influenza vaccination on numbers of US nursing home residents admitted to hospital: A cluster-randomised trial. The Lancet Respiratory Medicine 5(9), 738–746 (2017).

37. Jones. J. E., Sage. V. L. & Lakdawala. S. S. Viral and host heterogeneity and their effects on the viral life cycle. Nature Reviews Microbiology 19(4), 272–282(2020).

38. Kaiser. L., Fritz. R. S., Straus. S. E., Gubareva. L. & Hayden. F. G. Symptom pathogenesis during acute influenza: Interleukin-6 and other cytokine responses. Journal of Medical Virology 64(3), 262–268 (2001).

39. Hobson. D., Curry. R. L., Beare. A. S. & Ward-Gardner. A. The role of serum haemagglutination-inhibiting antibody in protection against challenge infection with influenza A2 and B viruses. Epidemiology and Infection 70(4), 767–777 (1972).

40. Palgen. J-L., et al. Integration of heterogeneous biological data in multiscale mechanistic model calibration: Application to lung adenocarcinoma. Acta Biotheoretica (2022).

41. Carrat. F., et al. Time lines of infection and disease in human influenza: A review of volunteer challenge studies. American Journal of Epidemiology 70(3), 19 (2008).

42. Sridharan. A., et al. Age-associated impaired plasmacytoid dendritic cell functions lead to decreased CD4 and CD8 T cell immunity. Age 33, 363–376 (2010).

43. Agrawal. A. & Gupta. S. Impact of aging on dendritic cell functions in humans. Ageing Research Reviews 10(3), 336–345 (2010).

44 Agrawal. A., Agrawal. S., Cao. J-N., Su. H., Osann. K. & Gupta. S. Altered innate immune functioning of dendritic cells in elderly humans: A role of phosphoinositide 3-kinase-signaling pathway. The Journal of Immunology 178(11), 6912–6922 (2014).

45 Lu. X., et al. Low quality antibody responses in critically ill patients hospitalized with pandemic influenza A(H1N1)pdm09 virus infection. Scientific Reports 12(1), 14971 (2022).

46. Treanor J. J. et al. Effectiveness of seasonal influenza vaccines in the United States during a season with circulation of all three vaccine strains. Clinical Infectious Diseases 55(7), 951–959 (2012).

47. Kniss. K., et al. Update: Influenza activity — United States, 2010–11 season, and composition of the 2011–12 influenza vaccine. Centers for Disease Control and Prevention: Morbidity and Mortality Weekly Report : 21 (2011)

48. Hadfield. J., et al. Nextstrain: Real-time tracking of pathogen evolution. Bioinformatics : 34(23), 4121–4123 (2018).

49. Cox. M. M. J., Patriarca. P. A. & Treanor. J. FluBlok, a recombinant hemagglutinin influenza vaccine. Influenza and Other Respiratory Viruses 2(6), 211–219 (2008).

50. Falsey. A. R., Treanor. J. J., Tornieporth. N., Capellan. J. & Gorse. G. J. Randomized, double-blind controlled phase 3 trial comparing the immunogenicity of high-dose and standard-dose influenza vaccine in adults 65 years of age and older. The Journal of Infectious Diseases 200(2), 172–180 (2009).

51. Allen. R., Rieger. T. & Musante. C. Efficient generation and selection of virtual populations in quantitative systems pharmacology models. CPT: Pharmacometrics & Systems Pharmacology 5(3), 140–146 (2016).

52. Rieger. T. R., et al. Improving the generation and selection of virtual populations in quantitative systems pharmacology models. Progress in Biophysics and Molecular Biology 139, 15–22 (2018).

53. Reed. C., et al. Estimates of the prevalence of pandemic (H1N1) 2009, United States, April–July 2009. Emerging Infectious Diseases 15(12), 2004 (2009).

54. Troeger, C. E. et al. Mortality, morbidity, and hospitalisations due to influenza lower respiratory tract infections, 2017: An analysis for the global burden of disease study 2017. The Lancet Respiratory Medicine 7(1), 69–89 (2019).

55. Lee, J. K. H. et al. Efficacy and effectiveness of high-dose versus standard-dose influenza vaccination for older adults: A systematic review and meta-analysis. Expert Review of Vaccines 17(5), 435–443 (2018).

56. Izurieta, H. S. et al. Relative effectiveness of cell-cultured and egg-based influenza vaccines among elderly persons in the United States, 2017–2018. The Journal of Infectious Diseases 220(8), 1255–1264 (2018).

57. Net, P., Colrat, F., Costa, M. N., Bianic, F., Thommes, E., Alvarez, F. P. Estimating public health and economic benefits along 10 years of fluzone® high dose in the United States. Vaccine 9, A56–A69 (2021).

58. Malosh, R. E., McGovern, I. & Monto, A. S. Influenza during the 2010–2020 decade in the United States: Seasonal outbreaks and vaccine interventions. Clinical Infectious Diseases 76(3), 540–549 (2022).

59. DiazGranados, C. A. et al. Efficacy of high-dose versus standard-dose influenza vaccine in older adults. New England Journal of Medicine 371(7), 635–645 (2014).

60. Borgonovo E, Plischke E. Sensitivity analysis: A review of recent advances. European Journal of Operational Research 248(3), 869–887 (2016).

61. Lee, H. Y. et al. Simulation and prediction of the adaptive immune response to influenza A virus infection. Journal of Virology 83(14), 7151–7165 (2009).

62. Dugan, H. L. et al. Preexisting immunity shapes distinct antibody landscapes after influenza virus infection and vaccination in humans. Science Translational Medicine 12(573), eabd3601 (2020).

63. Shay, D. K. et al. Comparative effectiveness of high-dose versus standard-dose influenza vaccines among US medicare beneficiaries in preventing postinfluenza deaths during 2012–2013 and 2013–2014. The Journal of Infectious Diseases 215(4), 510–517 (2017).

64. Robison, S. G. & Thomas, A. R. Assessing the effectiveness of high-dose influenza vaccine in preventing hospitalization among seniors, and observations on the limitations of effectiveness study design. Vaccine 36(45), 6683–6687 (2018).

65. Young-Xu, Y. et al. Relative vaccine effectiveness of high-dose versus standard-dose influenza vaccines among veterans health administration patients. The Journal of Infectious Diseases 217(11), 1718–1727 (2018).

66. Izurieta, H. S. et al. Relative effectiveness of influenza vaccines among the United States elderly, 2018–2019. The Journal of Infectious Diseases 22(2), 278–287 (2020).

67. Guthmiller, J. J., Utset, H. A. & Wilson, P. C. B cell responses against influenza viruses: Short-lived humoral immunity against a life-long threat. Viruses 13(6), 965 (2021).

68. Koroleva, M. et al. Heterologous viral protein interactions within licensed seasonal influenza virus vaccines. npj Vaccines 5(1), 3 (2020).

69. Einav, T., Gentles, L. E. & Bloom, J. D. SnapShot: Influenza by the numbers. Cell 182(2), 532–532 (2020).

70. Creskey, M. C. et al. Simultaneous quantification of the viral antigens hemagglutinin and neuraminidase in influenza vaccines by LC–MSE. Vaccine 30(32), 4762–4770 (2012).

71. Pollard, A. J. & Bijker, E. M. A guide to vaccinology: From basic principles to new developments. Nature Reviews Immunology 21(2), 83–100 (2020).

72. Ng, T. W. Y., Cowling, B. J., Gao, H. Z. & Thompson, M. G. Comparative immunogenicity of enhanced seasonal influenza vaccines in older adults: A systematic review and meta-analysis. The Journal of Infectious Diseases 219(10), 1525–1535 (2018).

73. Sridhar, S. et al. The potential of Beta variant containing COVID booster vaccines for chasing Omicron in 2022. Nature Communications 13(1), 5794 (2022).

74. Arsène, S. et al. Modeling the disruption of respiratory disease clinical trials by non-pharmaceutical COVID-19 interventions. Nature Communications 13(1), 1980 (2022).

75. Gálvez, J., Gálvez, J. J. & García-Peñarrubia, P. Is TCR/pMHC affinity a good estimate of the T-cell response? An answer based on predictions from 12 phenotypic models. Frontiers in Immunology 10, 349 (2019).

76. Linnik, J., Syedbasha, M., Hollenstein, Y., Halter, J., Egli, A. & Stelling, J. Model-based inference of neutralizing antibody avidities against influenza virus. PLOS Pathogens 18(1), e1010243 (2022).

77. Ross, T. M. et al. Influence of pre-existing hemagglutination inhibition titers against historical influenza strains on antibody response to inactivated trivalent influenza vaccine in adults 50–80 years of age. Human Vaccines & Immunotherapeutics 10(5), 1195–1203 (2014).

78. Hindmarsh, A. C. et al. SUNDIALS: Suite of nonlinear and differential/algebraic equation solvers. ACM Transactions on Mathematical Software (TOMS) 31(3), 363–396 (2005)

79. Lewis, N. M., Chung, J. R., Uyeki, T. M., Grohskopf, L., Ferdinands, J. M. & Patel, M. M. Interpretation of relative efficacy and effectiveness for influenza vaccines. Clinical Infectious Diseases 75(1), 170–175 (2021).

