## Supplementary information for "Multi-strain modeling of influenza vaccine effectiveness in older adults and its dependence on antigenic distance"

**This file includes:**

**Supplementary methods** 2

Supplementary Methods on submodels (Supplementary Figure S1) 2

Supplementary Methods on the multi-strain model 5

Simplifying hypotheses of the multi-strain model 5

Supplementary Methods on calibration 5

**Supplementary Figures** 8

Figure S1: Submodels 8

Figure S2: Calibration of virtual population 9

Figure S3: Contribution analysis on HI titer 11

**Supplementary tables** 12

Table S1: Variables 12

Table S2 : Values and references for the chosen fixed parameters 15

Table S3: sRVE against symptomatic infections, medical visits and hospitalization in the 65+ 15

**Also available in the supplementary materials are the SBML files for each of the submodels.**

### Supplementary methods

#### Supplementary Methods on submodels (Supplementary Figure S1)

*Immunization submodel* - Upon infection and vaccination, both innate and adaptive immune cells participate in the immune response to antigen encounter. We consider both of these responses, as well as naïve, activated, effector and memory cells, whose dynamics are described with mass action laws representing the transitions from these classes. The humoral response is central in the response against influenza infection and vaccination[1-2]. Upon antigen encounter, naïve B cells, with the the help of CD4[3-4], are known to enter a stage of somatic hypermutation and selection which leads to an increased fit between B cell receptors and antigens[5]. This affinity maturation is not modeled explicitly, but the newly activated B-cells are assumed to produce low avidity immunoglobulins G which neutralize viral antigens non-specifically[3]. B-cell activation is enhanced by the CD4 helper cells[4]. On a time scale of weeks to a month, activated B cells either go through apoptosis or produce memory B cells specific to the new antigen[5]. We do not distinguish between long-lived plasma cells and memory B cells[3], and we assume that, if the antigen is encountered a second time, simulated memory B cells will both boost the pre-existing memory B cells and produce high avidity antibodies that will neutralize the antigen more efficiently than the non-specific antibodies produced by activated B cell upon previous antigen encounter. Regarding the T-cell response, we consider only CD4+ and CD8+ cells. Upon presentation of the antigen, naïve CD4+ are activated and lead to the activation of several other cells (CD8+, B cells). On a time scale of days to a week, activated CD8+ cells target and kill infected cells by cytolysis[6]. On a time scale of weeks to a month, both CD4+ and CD8+ either go through apoptosis or are converted into memory T-cells (TRM, TEM and TCM cells) that will persist for a long time and do not need reactivation to perform their effector functions upon second antigen encounter[4, 7-11]. We disregard the process of positive and negative selection of T cells in the thymus. We simply assume that the cytolysis rate of CD8+ memory cells is higher if they are targeted against internal viral antigens than if they are targeted against HA and NA where their cytolysis rate is modulated by antigenic distances in the multi-strain model. We also consider NK cells that are activated by the presence of antigen and that kill infected cells by cytolysis with a rate which is lower than that of CD8+ memory cells at small antigenic distance. We disregard memory NK cells since their discovery is quite recent[12] and data is still sparse.

*Vaccine Immunogenicity submodel* - Split vaccines consist of immunogenic proteins which, in the case of influenza, have a molecular weight of about 60 kDa per monomer[13] . For this molecular weight, there is very little influx from tissue into systemic circulation[14]. Biodistribution data on particles with comparable size shows high accumulation in the lymph nodes themselves[15]. Therefore, antigen distribution beyond the lymph nodes is neglected. Given the molecular weight of HA and NA antigens (200 kDa/trimer for HA and 220 kDa/tetramer for NA[13]), it is assumed that these antigens can reach the lymph nodes by passive lymphatic drainage and by active transport after uptake by migratory antigen-presenting cells[14]. Although antigen-uptake is a dynamic process, we choose to model this by partitioning the initial antigen dose into direct and cell-based uptake and assume that this strategy is sufficient to generate realistic dynamics for mature dendritic cells in lymph nodes. The HD vaccine contains 4 times more hemagglutinin (60 μg against 15 μg) than the SD vaccine[16]. Split vaccines also contain detectable NA, about 0.1 μg per ug of hemagglutinin[17]. As the amount of NA is not standardized, we assume that HD contained 6 μg of NA while SD contained 1.5 μg, which corresponds to the relative amount of NA compared to HA that has been quantified in split vaccines[17] and in virions (300-400 HA trimers and 20-50 NA tetramers[13]). The HD vaccine was specifically designed to improve immune response and protection in older adults. Given the lack of data on the dependence of loaded dendritic cells on antigen dosage, we assume that a linear relationship is a sufficient approximation to the actual dose-response relationship. Furthermore, contributions to immunization from HA and NA are simply added without accounting for possible overlap, as dendritic cells express both. Effects of prior immunization on additional inflammation by immune complex recognition[18] or enhancement of dendritic cell functions[19] are not accounted for.

*Virus submodel* - Using a target-cell limited within-host model[20-27], the Influenza Virus Life Cycle submodel simulates variation in viral dynamics related to A/H1N1 and A/H3N2 subtypes. But in contrast to these classical within-host models, we do not simulate an eclipse phase as the delays in viral production (less than 12 hours) are usually lower than the one day time-step used in the multi-strain model. We assume that the HA-3 in A/H3N2 has a stronger affinity with the sialic acid epithelial cell receptor than the HA-1 in A/H1N1, resulting in partial immune escape in A/H3N2[28,29]. Viral replication rate is assumed to be lower in A/H3N2 infections than in A/H1N1 infections[30]. Hence, simulated A/H1N1 infections are more acute, but resolve generally faster than A/H3N2 infections[31]. Viral replication also happens at different rates for infectious and semi- or non-infectious viral particles so that the ratio of infectious to non-infectious viral particles decreases during infection with both subtypes[24,26]. The induced expression of Interferon Lambda 1 is known to vary according to viral subtypes and patients[32], so we assume that A/H3N2 inhibits antiviral host defenses more than A/H1N1 does, leading to a generally lower peak in antiviral cytokines in A/H3N2 infections in the VP. This makes the A/H3N2 infections more challenging to resolve.

*Pathogenesis submodel* - Using the Upper and Lower Respiratory Tract compartments, the pathogenesis submodel describes the effector response to viral exposure (neutralization, cytolysis, inflammation). All of these responses promote viral clearance but they may also contribute to tissue injury[33]. Specific neutralizing antibodies resulting from germinal centers have an avidity for HA and NA several orders of magnitude higher than non-specific antibodies which are present before an infection or vaccination is initiated[34], but a similar off rate binding. Due to lack of more specific data, the avidities for HA and NA have been set to the same values. The time evolution of specific neutralizing antibodies after affinity maturation (which is implicit in the model) is modeled in several compartments to simulate the response to infection (Upper/Lower Respiratory Tract, Blood) and to vaccination (Blood). Cytolysis of infected cells by CD8+ cells is modulated by antigenic distance if these cells target viral surface antigens but not when targeting viral internal antigens. Cytolysis of infected cells by NK cells is independent of antigenic distance. Regarding inflammation, IL-6 has been most closely associated with the development of symptoms and fever[35,36], so we assume that pro-inflammatory cytokines akin to IL-6 are produced in response to infection in the URT and LRT and that they define symptoms intensity. Viral shedding and symptoms are usually well correlated[31], with viral shedding peaking one day earlier than maximal symptoms. Viral clearance can take a few days in patients with mild disease but can take weeks in the URT and LRT of some patients with respiratory failure[35]. We disregarded infections that took more than one month to clear, as these patients were judged implausible or would have required additional treatment not modeled here. Pathogenesis is known to depend on patient age: immune responses are known to be poorer in the older population and most of the vaccines used in older adults offer limited protection or a limited duration of protection, particularly among those older than 75 years of age. Immunosenescence has been well documented and variation in disease severity and infection rate is mostly accountable by the variation in the host immune system. For instance, Sridharan et al. (2010) observed a severe decline in the production of IFN-III with age and proposed that it may explain the increased risk of influenza and other respiratory infections in the elderly38. Here, we assumed that age was associated with a decrease in the neutralization rate of antibodies[37], a decrease in the number of naïve cells[38], a decrease in antiviral cytokine autocatalysis (i.e. interferon type III)[38-40], and an increase in pro-inflammatory cytokine autocatalysis (i.e interleukin 6)[41].

#### Supplementary Methods on the multi-strain model

##### Simplifying hypotheses of the multi-strain model

Non-neutralizing antibodies are disregarded - Although the Matrix protein 2 (M2) is located at the viral membrane, antibodies are not commonly induced against M2 and it is currently unknown whether those rare antibodies contribute to protective humoral immunity[43]. Many reports have shown that antibodies against the Nuclear Protein (NP) are induced by natural infection and vaccination and are broadly reactive, however it remains controversial whether these non-neutralizing antibodies can protect against infection[43]. Non-neutralizing antibodies can also be generated against other internal proteins, including Matrix protein 1 (M1), the viral polymerases (PB1, PB2, and PA), Nuclear Export Protein (NEP), and Non-Structural protein 1 (NS1). However, the general sero-prevalence of antibodies against these epitopes is not well defined[43]. The general quantity of these internal proteins, with the exception of M1, is considerably lower than that of HA, NA, and NP and, therefore, antibodies may be rarely induced against these antigens anyway. In our model, we disregard all these subdominant non-neutralizing antibodies, due to lack of data and consensus on their role in infection or vaccination.

Antibodies against HA stalk are disregarded - It is known that, if the antigenic distance between the new viral exposure and previously encountered viruses is very large (i.e. several decades apart), people will preferentially recall broadly reactive memory B cells, usually against HA-stalk, as was the case for first exposure to the A/H1N1pdm09 virus in 2009 in the elderly population[43]. However, as the antigenic distance between viruses is small (i.e. less than a decade apart), people rather tend to recall memory B cells against variable epitopes of the HA head[43]. We disregard imprinting from childhood and broadly neutralizing antibodies against HA-stalk to concentrate our simulations on seasonal variations over a relatively short time span.

From studies of individuals with inherited or acquired immunodeficiency, it is clear that whereas antibody deficiency increases susceptibility to acquisition of infection, T cell deficiency results in failure to control a pathogen after infection[44,45]. Although evidence for the involvement of T cells in vaccine-induced protection is limited, this is likely owing, in part, to difficulties in accessing T cells to study as only the blood is easily accessible, whereas many T cells are resident in tissues such as lymph nodes[44]. Our model is phenomenological in aspects that relate to the relationship between the affinity of T cell receptor (TCR) and the effector functions of T cells (cytolysis, helper function). Despite extensive experimental work on TCR affinity, it has not been possible to establish a clear correlation between affinity and T-cell response because the available data are far from conclusive and even contradictory[46]. So the response of T-cells targeted at viral surface antigens (HA, NA) is just modulated by antigenic distance, but the response of T-cells targeted at internal antigens is not.

#### Supplementary Methods on calibration

We summarize the different calibration steps undertaken as follows.

Steps 1 to 5 correspond to the calibration of a single parameterization to derive a reference vaccine, two reference A viral subtypes and 5 reference patients exhibiting various degrees of disease severity and response to vaccination. Steps 6 to 10 correspond to calibration at the population level, where the goal is to derive a cohort of virtual patients exhibiting the known frequencies of clinical outcomes in different clinical arms (control versus vaccine arms) or differences between age groups (i.e. decrease of treatment efficacy with age).

**Step 1:** Using mouse biodistribution and lymph flow data[47-49], we estimate the relative quantity and dynamics of antigens arriving at lymph nodes by direct lymph drainage and by APC uptake. To calibrate realistic APC dynamics, we use timeseries of the number of primed APCs in non-human primates in response to administration of a reference vaccine dose of 100 µg of fluorescent HIV-1 envelope glycoprotein Env[18, 50].

**Step 2:** We calibrate the temporal evolution of infectious (TCID50/mL) and non-infectious viral load (RNA copies/mL) quantified in A549 cells infected *in vitro* by A/(H1N1) pdm09 at a given multiplicity of infection (moi)[24, 26].

**Step 3:** We then calibrate the temporal evolution of infectious viral load (TCID50/mL) and IFN-type III secretion quantified in Normal Human Bronchial Epithelial cells (NHBE) from 2 different donors, infected *in vitro* by 2 reference strains of different subtypes: A/(H1N1)pdm09 and A/H3N2/Perth[32]. This data allows the calibration of the variability between subtypes. As the main variability in viral load is due to the subtype we consider the parameters tied to the replicative kinetics to vary only according to the subtype (and not the strain), and stay fixed across the virtual population, whereas the cytokine secretion and intensity of antiviral action varies among patients.

**Step 4:** As the adaptive immune cell activation peaks at a similar time in both mice and humans, at about 10 days after infection[1,6], we calibrate the temporal evolution of the immune system in response to infection using timeseries derived from *in vivo* studies in mice[5, 51, 52].

Typically, the broad initial response was followed by a period of titer decay during which antibody titers stabilized to form an altered antibody landscape over the course of ~1 year[53]. Antibody responses were first detected between 1 and 17 days post-infection, which is consistent with previous reports of antibody responses being detected 7 days post-vaccination.

**Step 5:** We define 5 reference patients (Fig. 2). Reference patients 1, 2 and 3 exhibit respectively an asymptomatic, a mild symptomatic or a severe symptomatic infection in absence of vaccination, and no breakthrough infection after vaccination. Reference patients 4 and 5 exhibit vaccine breakthrough infection, with and without achieving seroprotective titers 28 days after vaccination respectively. These 5 reference patients are used to define the initial virtual population (Supplementary Fig. S2).

Patients are selected such that in the control arm, they develop an asymptomatic or symptomatic infection after a single exposure, which occurs at a variable time during the flu season, between December and April, with a peak incidence around end of January - beginning of February. Each virtual patient is his own control, so the same patient is exposed to the same infectious quantity and concentration of the same virus at the same moment in the control and the two vaccine arms.

**Step 6:** We adapt the distributions of patient descriptors to reproduce the variability in symptoms and disease severity after exposure to influenza A virus in non-naïve human population[31].

**Step 7:** We adapt the correlations between some patient descriptors to reproduce immunosenescence in the VP, according to the estimations reported by the CDC[54]. These estimations are based on A/H1N1pdm09 study and are possibly underestimating the average hospitalization rate in people born before 1950s, as they were less at risk of severe infection by the A/H1N1pdm09 due to childhood imprinting patterns[43], so we use these estimations as a lower bound only.

**Step 8:** We adapt the baseline patient descriptors to reproduce a plausible variability in patient immune history. As the kinetics of humoral immune response differ in vaccinees immunized for the first time from those who had been vaccinated previously[55], we assume virtual patients are vaccinated for the first time. We calibrate the residual immunity of those previously unvaccinated patients so that the geometric mean HI titers (tested against historical strains) of the virtual population is inferior to 40, as was reported against both A subtypes in adults older than 50 years old[56].

**Step 9:** We calibrate the range of humoral response after infection and vaccination using the physiological range of specific IgG in serum ([0, 420] microg/mL[57]. In our model, this range corresponds to log2 HI titers in the range 1-14, with 1 being the minimal experimental value corresponding to dilution 1:5 and 14 corresponding to dilutions 1:40960.

**Step 10:** We sample the patients of the VP to better match the seroprotection rate reported in randomized clinical trials[58,59] and the average proportion of prevented symptomatic infections reported in RWD during the 2010-2011 season with low antigenic match between vaccine and circulating strain, where Treanor et al. (2012) estimated an overall adjusted VE of 60 %[60]. The population sampling is done using algorithms similar to those reported previously[61].

We aimed at having at most 30% of patients aged more than 50 years not mounting a robust humoral response after vaccination with splitSD[58, 59]. We calibrated the HI titers against historical strains to be below the seroprotection threshold for the vast majority of our older patients, so that most of the virtual patients that fail to reach seroprotection levels against the vaccine strain at 28 days post-vaccination is due to immunosenescence rather than already built protection thanks to strong pre-existing humoral or cellular immune response (although we cannot exclude this can occur).

#### Supplementary Figures

##### Figure S1: Submodels

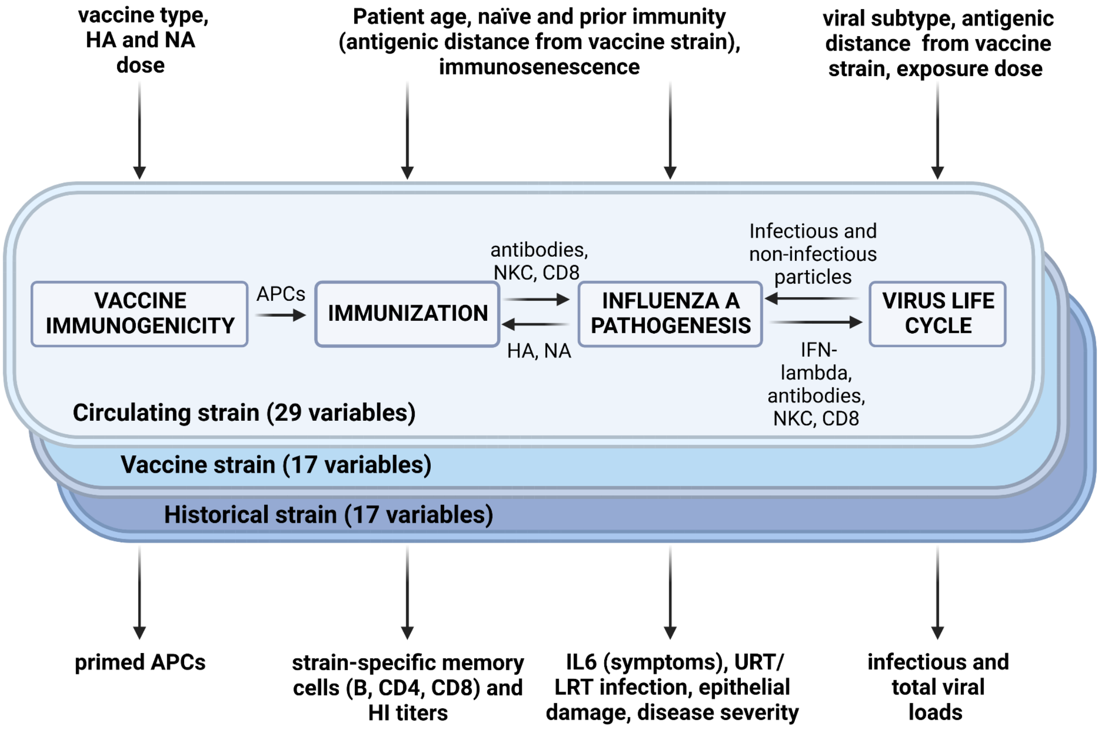


**Legend**: Submodel connections and multiplications to simulate immunity specific to historical strains, vaccine strains and circulating strains. Top: inputs of the multi-strain combination of the submodels. Bottom: outputs of the multi-strain combination of the submodels. APCs: antigen-presenting cells, NKC: Natural killer cells. B, CD4 and CD8 cells are simulated in different states (naïve, primed, activated, effector, memory). HA: hemagglutinin viral surface antigen. NA: neuraminidase viral surface antigen. IL-6: interleukin 6 with pro-inflammatory properties. IFN-lambda: interferon of type III with antiviral properties.

#### Figure S2: *Calibration of virtual population.*

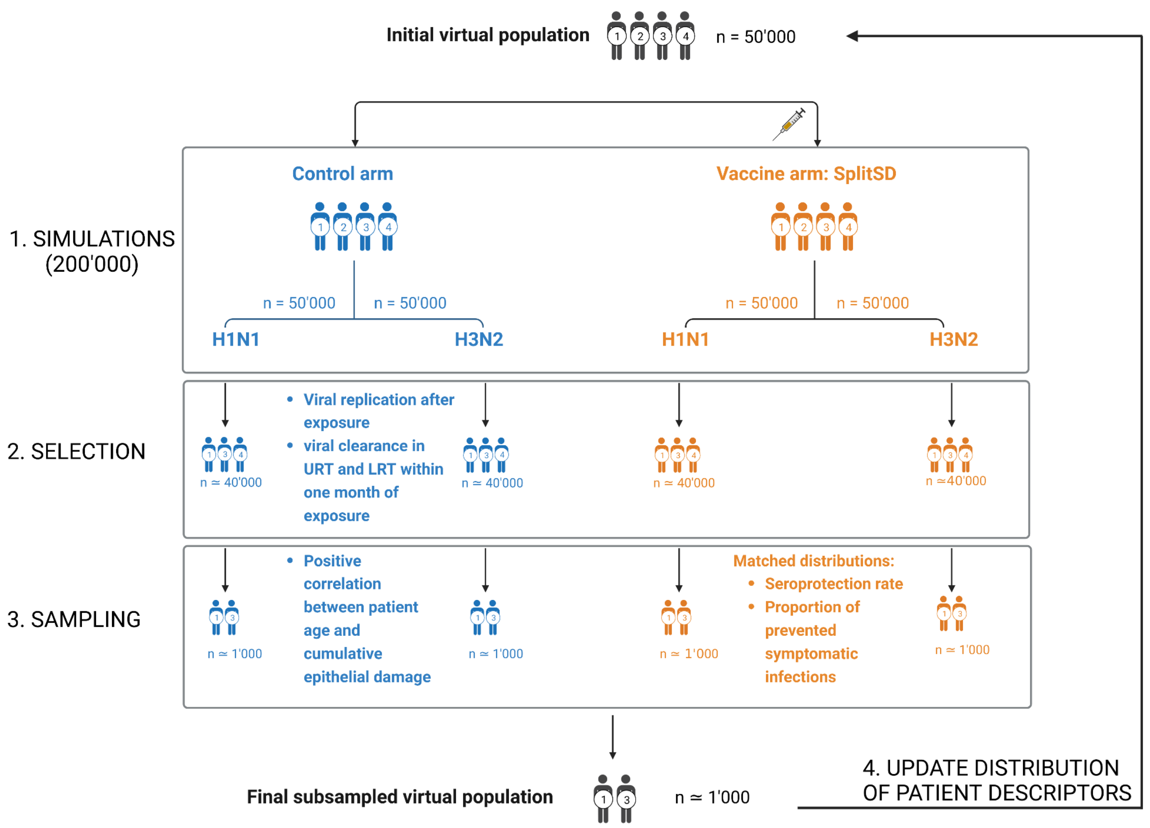


**Legend**: Scheme presenting the different steps to calibrate the virtual population following the subsampling method described in Allen et al. (2016)[61] and Rieger et al. (2018)[62]. After having defined plausible distributions of patient descriptors (about 60 parameters) and their putative correlations (i.e. with age), a large plausible virtual population (50'000 patients) is simulated in four clinical arms, representing a control and a treatment arm where the split standard dose vaccine is administered or not at day 1, followed by a viral exposure to H1N1 or H3N2 occurring during the epidemic season 2010-2011. The time of exposure depends on the patient (not on the viral subtype), and is normally distributed within the population with an average of 90 days, a standard deviation of 20 days, ranging between day 15 and day 150. This results in 200'000 simulations (step 1), where each patient is simulated in 4 scenarios. In a second step, the plausible patients are selected based on the infection characteristics in the control group: only patients exhibiting viral replication after exposure as well viral clearance in the Upper and Lower Respiratory Tract (URT/LRT) within one month are selected. This step avoids parameter combinations which lead to either too strong or too weak immunity against infection in the control arm, as these patients would be uninformative (always protected), rare or unlikely (unable to clear an infection within a reasonable time). In a third step, the selected plausible patients (~40'000) are assigned a probability of inclusion in the virtual population based on their ability to conjointly match the clinical distributions of proportion of prevented symptomatic infections due to vaccine administration and seroprotection rates (after vaccination) for both viral subtypes which circulated similarly in the USA. The patients are also sampled in order to generate a positive correlation between age and cumulative epithelial damage after infection in the control group, in order to emulate the increased risk of hospitalization with age. In a fourth step, the distribution of patient descriptors is updated to increase the probabilities of patients to be included in the virtual population and the whole procedure is performed again and again until a sufficient number of unique patients (~1'000) is found that match the objectives of immunosenescence, seroprotection rate and proportion of prevented symptomatic infections sufficiently well (Table 2).

#### Figure S3: Contribution analysis on HI titer

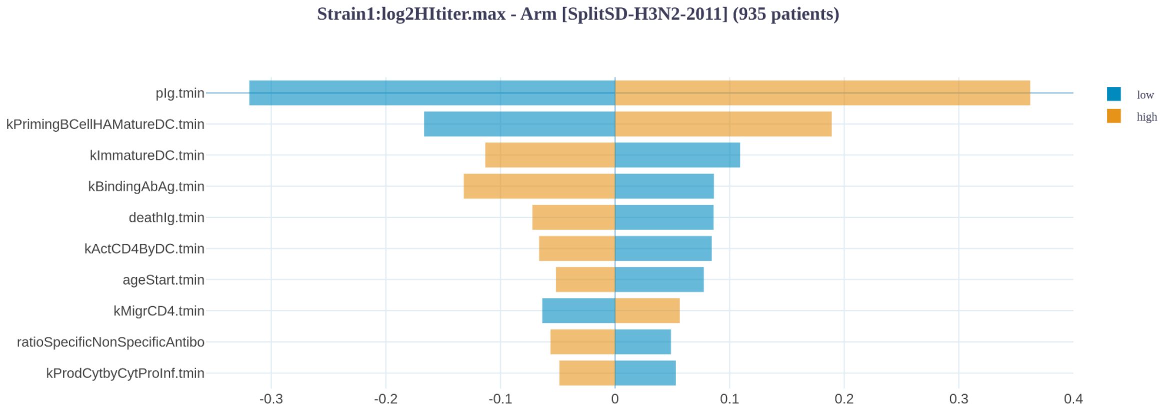


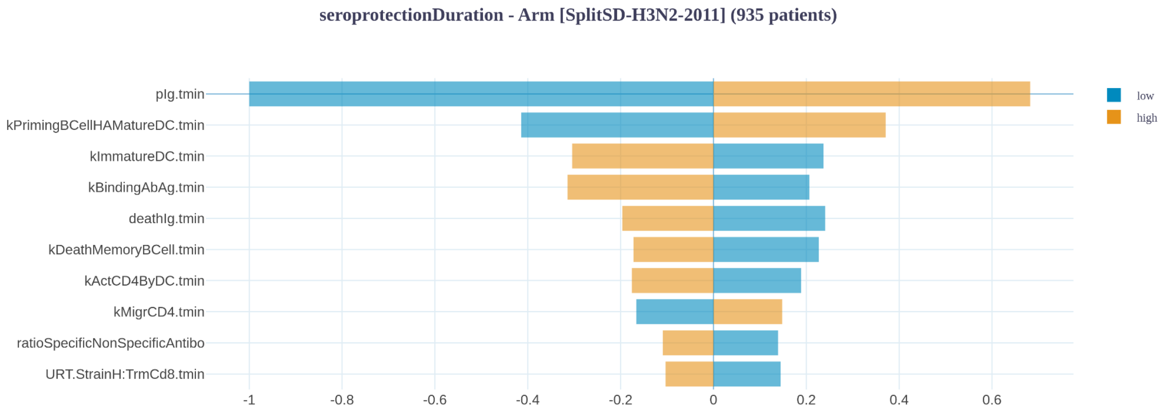


Contribution analysis on maximum of HI titer (top) and seroprotection levels (bottom), i.e. the quantity of time that the patient spent with a HI titer above 4. A positive correlation between a marker of response to vaccination (i.e. HI titer) is signified by low (blue) to high (orange) values from left to right, while a negative correlation goes from high (orange) to low (blue) values from left to right. Values are expressed in % change of the subpopulation’s median compared to the whole population’s median.

### Supplementary tables

#### Table S1: Variables

| Name | Definition | Compartment | Strain-specific |
|---|---|---|---|
| hEC | Population of susceptible healthy epithelial cells in the lungs | URT, LRT | No |
| iEC | Population of infected cells | URT, LRT | Yes |
| immatureDC | Population of immature dendritic cells | URT, LRT, Injection site | No |
| matureDC | Population of mature dendritic cells (antigen-presenting cells) | Lymph nodes*, URT, LRT, Injection site, Blood | Yes |
| virusLoadedDC | Virus loaded dendritic cells in the blood | Lymph nodes*, URT, LRT, Injection site, Blood | Yes |
| NaiveBCell | Population of naive B cells | Lymph nodes* | No |
| PrimedBCell | Population of primed B cells by antigen-presenting cells | Lymph nodes* | Yes |
| ActivatedBCell | Population of activated B cells (by direct antigen binding) | URT, LRT | Yes |
| MemoryBCell | Population of memory B cells | Lymph nodes*, Blood | Yes |

| | | | |
|---|---|---|---|
| NaiveCD4 | Population of naive CD4 cells | Lymph nodes* | No |
| EffectorCD4 | Population of effector CD4 cells | Lymph nodes*, URT, LRT | Yes |
| TrmCD4 | Population of tissue resident CD4 cells (Trm CD4) | URT, LRT | Yes |
| TemCd4 | Population of effector memory CD4 cells (Tem CD4) | Blood | Yes |
| TcmCd4 | Population of central memory CD4 cells (Tcm CD4) | Lymph nodes* | Yes |
| NaiveCD8 | Population of naive CD8 cells | Lymph nodes* | No |
| EffectorCD8 | Population of cytotoxic CD8 cells | Lymph nodes* | Yes |
| TrmCd8 | Population of tissue resident CD8 cells (Trm CD8) | URT, LRT | Yes |
| TemCd8 | Population of effector memory CD8 cells (Tem CD8) | Blood | Yes |
| TcmCd8 | Population of central memory CD8 cells (Tcm CD8) | Lymph nodes* | Yes |
| ActivatedNKC | Population of activated natural killer cells (NKC) | Blood | No |
| NKC | Population of natural killer cells (NKC) | Blood | No |
| tCL | Cytotoxic T cells | URT, LRT | Yes |
| IgNS | Concentration of non-specific antibodies | URT, LRT | No |

| | | | |
|---|---|---|---|
| Ig | Concentration of specific antibodies | Blood, URT, LRT | Yes |
| ProInfCyt | Concentration of type I cytokines (pro-inflammatory, IL6) | URT, LRT | No |
| AntiviralCyt | Concentration of type III cytokines (antiviral, IFNlambda) | URT, LRT | No |
| InfectiousIV | Infectious viral load in the extracellular space of the lungs | URT, LRT | Yes |
| NonInfectiousIV | Non-infectious viral load in the extracellular space of the lungs | URT, LRT | Yes |
| TotIV | Total viral load (infectious + non-infectious) | URT, LRT | Yes |
| ivHA | Concentration of HA antigen from viral infection | URT, LRT, Lymph nodes*, Blood, injection site (Muscle) | Yes |
| ivNA | Concentration of NA antigen from viral infection | URT, LRT, Lymph nodes*, Blood, injection site (Muscle) | Yes |
| HAdose | Concentration of HA from vaccination (depends on vaccine type) | Injection site | Yes |
| NAdose | Concentration of NA from vaccination (depends on vaccine type) | Injection site | Yes |

* “Lymph nodes” are actually several compartments: Lymph nodes tied to URT, LRT and injection site (Muscle).

#### Table S2 : Values and references for the chosen fixed parameters.

| Fixed parameters | Description | Reference average value |
|---|---|---|
| kAvidityHAspec* | Avidity of specific neutralizing antibodies for HA antigen | 1e9 / M (= 1 L/nmol) as the average of 1 / KD in Figure 5C[34] and in Figure S4C of Horns et al., 2020[34]. It might be reasonable to assume a lower KD for H1 than for H3 (1e-10 M instead of 1e-9 M; meaning a higher avidity 1e10 / M instead of 1e9 / M for H1) but this difference has not been taken into account in the model so far. |
| kAvidityHAnonspec | Avidity of non-specific neutralizing antibodies for HA antigen | Vaccinated individuals have antibodies that bind the virus protein $10^2$ to $10^3$ times as strongly[42]. From this assertion, we assumed that a reasonable value for this parameter was 1e7 / M (= 0.01 L/nMol). |
| kAvidityNAspec | Avidity of specific neutralizing antibodies for NA antigen | No data found, so it was assumed that kAvidityNAspec = kAvidityHAspec = 1e9 / M |
| kAvidityNAnonspec | Avidity of specific neutralizing antibodies for NA antigen | No data found, so it was assumed that kAvidityNAnonspec = kAvidityHAnonspec = 1e7 / M |

**Legend: The * marks parameters which directly affect HI titers as tested in experimental assays.**

##### Table S3: sRVE against symptomatic infections, medical visits and hospitalization in the 65+.

| INPUTS | | | | | | OUTPUTS | | |
|---|---|---|---|---|---|---|---|---|
| Season | Age class | Number of virtual patients | Main subtype #1 | Antigenic distance HA #2 | Antigenic distance NA #2 | Predicted RVE (symptomatic infections) | Predicted RVE (medical visit) | Predicted RVE (hospitalization) |
| 2011-2012 | 65-96 | 622 | H3N2 | 0.077 | 0 | 7 | 2 | 48 |
| 2012-2013 | 65-96 | 622 | H3N2 | 0.00458 | 0.00702 | 15 | 9 | 69 |
| 2013-2014 | 65-96 | 622 | H1N1 | 0 | 0.0123 | 19 | 13 | 57 |
| 2014-2015 | 65-96 | 622 | H3N2 | 0.116 | 0 | 7 | 0 | 43 |
| 2015-2016 | 65-96 | 622 | H1N1 | 0 | 0.0141 | 19 | 15 | 52 |
| 2016-2017 | 65-96 | 622 | H3N2 | 0 | 0.109 | 14 | 8 | 65 |
| 2017-2018 | 65-96 | 622 | H3N2 | 0.0466 | 0.223 | 9 | 3 | 49 |

| | | | | | | | | |
|---|---|---|---|---|---|---|---|---|
| **2018-2019** | 65-96 | 622 | H1N1 | 0.0127 | 0 | 20 | 15 | 48 |
| **2018-2019** | 65-96 | 622 | H3N2 | 0.15 | 0.0573 | 4 | -4 | 44 |
| **2019-2020** | 65-96 | 622 | H1N1 | 0.0892 | 0 | 14 | 13 | 17 |
| **2021-2022** | 65-96 | 622 | H3N2 | 0.461 | 0.113 | 2 | 2 | 5 |

References

1. Lam J.H., Baumgarth N. The multifaceted b cell response to influenza virus. *J Immunol* **202(2)**, 351–359 (2019)

2. Rodda L.B., Pepper M. Both naive and memory b cells respond to flu vaccine. *Nature* **586**, 34-35 (2020).

3. Akkaya M., Kwak K., Pierce S.K. B cell memory: Building two walls of protection against pathogens. *Nature Reviews Immunology* **20**, 229–238 (2020).

4. MacLeod M.K.L., Kappler J.W., Marrack P. Memory CD4 t cells: Generation, reactivation and re-assignment. *Immunology* **130**, 10-15 (2010).

5. Yewdell W.T. et al. Temporal dynamics of persistent germinal centers and memory b cell differentiation following respiratory virus infection. *Cell reports* **37**, no. 6 (2021).

6. Schmidt M.E., Varga S.M. The CD8 t cell response to respiratory virus infections. *Frontiers in Immunology* **9**, 678 (2018).

7. Cho B.K., Wang C., Sugawa S., Eisen H.N., Chen J. Functional differences between memory and naive CD8 t cells. *Proceedings of the National Academy of Sciences* **96**, 2976-2981 (1999).

8. Woodland D.L., Kohlmeier J.E. Migration, maintenance and recall of memory t cells in peripheral tissues. *Nature Reviews Immunology* **9**, 153–161 (2009).

9. Mueller S.N., Gebhardt T., Carbone F.R., Heath W.R. Memory t cell subsets, migration patterns, and tissue residence. *Annual Review of Immunology* **31**, 137-161 (2012).

10. Wu X., Wu P., Shen Y., Jiang X., Xu F. CD8+ resident memory t cells and viral infection. *Frontiers in Immunology* **9** (2018).

11. Raphael I., Joern R.R., Forsthuber T.G. Memory CD4+ t cells in immunity and autoimmune diseases. *Cells* **9(3)**, 531 (2020).

12. Dou Y. et al. Influenza vaccine induces intracellular immune memory of human NK cells. *PloS one*, **10 (3)** (2015).

13. Einav T., Gentles L.E., Bloom J.D. SnapShot: Influenza by the numbers. *Cell*, **182(2)**, 532-532 (2020).

14. Moyer T.J., Zmolek A.C., Irvine D.J. Beyond antigens and adjuvants: Formulating future vaccines. *Journal of Clinical Investigation*, **126(3)**, 799-808 (2016).

15. Katagiri W. et al. Real-time imaging of vaccine biodistribution using zwitterionic NIR nanoparticles. *Advanced Healthcare Materials*, **8 (15),** 1900035 (2019).

16. Gravenstein S. et al. Comparative effectiveness of high-dose versus standard-dose influenza vaccination on numbers of US nursing home residents admitted to hospital: A cluster-randomised trial. *The Lancet Respiratory Medicine*, **5(9)**, 738-746 (2017).

17. Koroleva M. et al. Heterologous viral protein interactions within licensed seasonal influenza virus vaccines. *npj Vaccines*, **5(1)**, 3 (2020).

18. Ols S. et al. Route of vaccine administration alters antigen trafficking but not innate or adaptive immunity. *Cell Reports* **30(12)**, 3964-3971 (2020).

19. Lu L.L., Suscovich T.J., Fortune S.M., Alter G. Beyond binding: Antibody effector functions in infectious diseases. *Nature Reviews Immunology* **18(1)**, 46-61 (2017).

20. Baccam P., Beauchemin C., Macken C.A., Hayden F.G., Perelson A.S. Kinetics of influenza a virus infection in humans. *Journal of Virology* **80(15)**, 7590-7599 (2006).

21. Saenz R.A. et al. Dynamics of influenza virus infection and pathology. *Journal of Virology* **84(8)**, 3974-3983 (2010).

22. Pawelek K.A., Huynh G.T., Quinlivan M., Cullinane A., Rong L., Perelson A.S. Modeling within-host dynamics of influenza virus infection including immune responses. *PLoS Computational Biology* **8(6)**, 1002588 (2012).

23. Li Z., Zhou H., Lu Y., Colatsky T. A critical role for immune system response in mediating anti-influenza drug synergies assessed by mechanistic modeling. *CPT: Pharmacometrics & Systems Pharmacology* **3(0)**, 1-9 (2014).

24. Simon P.F. et al. Avian influenza viruses that cause highly virulent infections in humans exhibit distinct replicative properties in contrast to human H1N1 viruses. *Scientific Reports*, **6(1)**, 24154 (2016).

25. Zarnitsyna V.I., Handel A., McMaster S.R., Hayward S.L., Kohlmeier J.E., Antia R. Mathematical model reveals the role of memory CD8 t cell populations in recall responses to influenza. *Frontiers in Immunology* **7**, 165 (2016).

26. Yan A.W.C., Zhou J., Beauchemin C.A.A., Russell C.A., Barclay W.S., Riley S. Quantifying mechanistic traits of influenza viral dynamics using in vitro data. *Epidemics* **33**, 100406 (2020).

27. Samieegohar M. et al. Calibration and validation of a mechanistic COVID-19 model for translational quantitative systems pharmacology – a proof-of-concept model development for remdesivir. *Clinical Pharmacology & Therapeutics* **112(4),** 882-891 (2022).

28. Hensley S.E. et al. Hemagglutinin receptor binding avidity drives influenza a virus antigenic drift. *Science* **326(5953)**, 734-736 (2009).

29. Li Y. et al. Single hemagglutinin mutations that alter both antigenicity and receptor binding avidity influence influenza virus antigenic clustering. *Journal of Virology* **87(17)**, 9904-9910 (2013).

30. Mitchell H., Levin D., Forrest S., Beauchemin C.A., Tipper J., Knight J., Donart N., Layton R.C., Pyles J., Gao P., Harrod K.S. Higher level of replication efficiency of 2009 (H1N1) pandemic influenza virus than those of seasonal and avian strains: Kinetics from epithelial cell culture and computational modeling. *Journal of virology* **85(2)**, 1125-35 (2011).

31. Carrat F. et al. Time lines of infection and disease in human influenza: A review of volunteer challenge studies. *American Journal of Epidemiology* **167(7)**, 775-785 (2008).

32. Ilyushina N.A., Dickensheets H., Donnelly R.P. A comparison of interferon gene expression induced by influenza A virus infection of human airway epithelial cells from two different donors. *Virus Research* **264**, 1-7 (2019).

33. Herold S., Becker C., Ridge K.M., Budinger G.R.S. Influenza virus-induced lung injury: Pathogenesis and implications for treatment. *European Respiratory Journal* **45(5)**, 1463-1478 (2015).

34. Horns F., Dekker C.L., Quake S.R. Extended: Memory b cell activation, broad anti-influenza antibodies, and bystander activation revealed by single-cell transcriptomics. *Cell Reports* **30(3)**, 905-913 (2020).

35. Hayden F.G., Fritz R., Lobo M.C., Alvord W., Strober W., Straus S.E. Local and systemic cytokine responses during experimental human influenza a virus infection. Relation to

symptom formation and host defense. *Journal of Clinical Investigation* **101(3)**, 643-649 (2008).

36. Kaiser L., Fritz R.S., Straus S.E., Gubareva L., Hayden F.G. Symptom pathogenesis during acute influenza: Interleukin-6 and other cytokine responses. *Journal of Medical Virology* **64(3)**, 262-268 (2001).

37. Lu X. et al. Low quality antibody responses in critically ill patients hospitalized with pandemic influenza a(H1N1)pdm09 virus infection. *Scientific Reports* **12(1)**, 14971 (2022).

38. Sridharan A. et al. Age-associated impaired plasmacytoid dendritic cell functions lead to decreased CD4 and CD8 t cell immunity. *Age* **33**, 363-376 (2010).

39. Agrawal A., Gupta S. Impact of aging on dendritic cell functions in humans. *Ageing Research Reviews* **10(3)**, 336-345 (2010).

40. Agrawal A., Agrawal S., Cao J-N., Su H., Osann K., Gupta S. Altered innate immune functioning of dendritic cells in elderly humans: A role of phosphoinositide 3-kinase-signaling pathway. *The Journal of Immunology* **178(11)**, 6912-6922 (2014).

41. Panda A. et al. Age-associated decrease in TLR function in primary human dendritic cells predicts influenza vaccine response. *The Journal of Immunology* **184(5)**, 2518-2527 (2010).

42. Deem M.W., Hejazi P. Theoretical aspects of immunity. *Annual Review of Chemical and Biomolecular Engineering* **1**, 247-276 (2010).

43. Guthmiller J.J., Utset H.A., Wilson P.C. B cell responses against influenza viruses: Short-lived humoral immunity against a life-long threat. *Viruses* **13(6)**, 965 (2021).

44. Pollard A.J., Bijker E.M. A guide to vaccinology: From basic principles to new developments. *Nature Reviews Immunology* **21(2)**, 83-100 (2020).

45. L'Huillier A.G. et al. T-cell responses following natural influenza infection or vaccination in solid organ transplant recipients. *Scientific Reports* **10(1)**, 10104 (2020).

46. Gálvez J., Gálvez J.J., García-Peñarrubia P. Is TCR/pMHC affinity a good estimate of the t-cell response? An answer based on predictions from 12 phenotypic models. *Frontiers in Immunology* **10**, 349 (2019).

47. Gerner M.Y., Torabi-Parizi P., Germain R.N. Strategically localized dendritic cells promote rapid t cell responses to lymph-borne particulate antigens. *Immunity* **42(1)**, 172-185 (2014).

48. Woodruff M.C. et al. Trans-nodal migration of resident dendritic cells into medullary interfollicular regions initiates immunity to influenza vaccine. *Journal of Experimental Medicine* **211(8)**, 1611-1621 (2014).

49. Winde C.M. de, Munday C., Acton S.E. Molecular mechanisms of dendritic cell migration in immunity and cancer. *Medical Microbiology and Immunology* **209(4)**, 515-529 (2020)

50. Liang F. et al. Vaccine priming is restricted to draining lymph nodes and controlled by adjuvant-mediated antigen uptake. *Science Translational Medicine* **9(393)**, eaal2094 (2017).

51. Lawrence C.W., Ream R.M., Braciale T.J. Frequency, specificity, and sites of expansion of CD8+t cells during primary pulmonary influenza virus infection. *The Journal of Immunology* **174(9)**, 5332-5340 (2005).

52. Robinson A.M., Higgins B.W., Shuparski A.G., Miller K.B., McHeyzer-Williams L.J., McHeyzer-Williams M.G. Evolution of antigen-specific follicular helper t cell transcription from effector function to memory. *Science Immunology* **7(76)**, eabm2084 (2022).

53. Fonville J.M., Wilks S.H., James S.L., Fox A., Ventresca M., Aban M., Xue L., Jones T.C., Le N.M., Pham Q.T., Tran N.D. Antibody landscapes after influenza virus infection or vaccination. *Science* **346(6212)**, 996-1000 (2014).

54. Reed C. et al. Estimates of the prevalence of pandemic (H1N1) 2009, united states, april–july 2009. *Emerging Infectious Diseases* **15(12)**, 2004 (2009).

55. Künzel W. Kinetics of humoral antibody response to trivalent inactivated split influenza vaccine in subjects previously vaccinated or vaccinated for the first time. *Vaccine* **14(12)**, 1108-1110 (2002).

56. Hallmann-Szelińska E., Szymański K., Łuniewska K., Kondratiuk K., Brydak L.B. Hemagglutination inhibition antibody titers as a correlate of protection against influenza disease in the 2018/2019 epidemic season in poland. *Acta Biochimica Polonica* **67(1)**, 93-98 (2020).

57. Linnik J., Syedbasha M., Hollenstein Y., Halter J., Egli A., Stelling J. Model-based inference of neutralizing antibody avidities against influenza virus. *PLOS Pathogens* **18(1)**, e1010243 (2022).

58. Cox M.M.J., Patriarca P.A., Treanor J. FluBlok, a recombinant hemagglutinin influenza vaccine. *Influenza and Other Respiratory Viruses* **2(6)**, 211-219 (2008)

59. Falsey A.R., Treanor J.J., Tornieporth N., Capellan J., Gorse G.J. Randomized, double-blind controlled phase 3 trial comparing the immunogenicity of high-dose and standard-dose influenza vaccine in adults 65 years of age and older. *The Journal of Infectious Diseases* **200(2)**, 172-180 (2009).

60. Treanor J.J. et al. Effectiveness of seasonal influenza vaccines in the united states during a season with circulation of all three vaccine strains. *Clinical Infectious Diseases* **55(7)**, 951-959 (2012).

61. Allen R., Rieger T., Musante C. Efficient generation and selection of virtual populations in quantitative systems pharmacology models. *CPT: Pharmacometrics & Systems Pharmacology* **5(3)**, 140-146 (2016).

62. Rieger T.R., Allen R.J., Bystricky L., Chen Y., Colopy G.W., Cui Y., Gonzalez A., Liu Y., White R.D., Everett R.A., Banks H.T. Improving the generation and selection of virtual populations in quantitative systems pharmacology models. *Progress in biophysics and molecular biology* **139**, 15-22 (2018).
